# Multi-cohort analysis of host immune response identifies conserved protective and detrimental modules associated with severity irrespective of virus

**DOI:** 10.1101/2020.10.02.20205880

**Authors:** Hong Zheng, Aditya M Rao, Denis Dermadi, Jiaying Toh, Lara Murphy Jones, Michele Donato, Yiran Liu, Yapeng Su, Minas Karagiannis, Theodoros Marantos, Yehudit Hasin-Brumshtein, Yudong D He, Evangelos J Giamarellos-Bourboulis, Jim Heath, Purvesh Khatri

## Abstract

SARS-CoV-2 pandemic, the fourth pandemic of the decade, has underscored gaps in global pandemic preparedness and the need for generalizable tests to avert overwhelming healthcare systems worldwide, irrespective of a virus. We integrated 4,780 blood transcriptome profiles from patients infected with one of 16 viruses across 34 independent cohorts from 18 countries, and 71 scRNA-seq profiles of 264,224 immune cells across three independent cohorts. We found a myeloid cell-dominated conserved host response associated with severity. It showed increased hematopoiesis, myelopoiesis, and myeloid-derived suppressor cells with increased severity. We identified four gene modules that delineate distinct trajectories associated with mild and severe outcomes, and show the interferon response was decoupled from protective host response during severe viral infection. These modules distinguished non-severe from severe viral infection with clinically useful accuracy. Together, our findings provide insights into immune response dynamics during viral infection, and identify factors that may influence patient outcomes.

## Introduction

Outbreaks of infectious diseases globally have been increasing steadily over the last 40 years (Christiansen, 2018). The first two decades of the 21^st^ century have been marked by seven outbreaks of novel viral infections that include severe acute respiratory syndrome coronavirus (SARS-CoV-1; 2002), H1N1 influenza (2009), Middle East Respiratory Syndrome Coronavirus (MERS-CoV; 2012), chikungunya (2014), Ebola (2014), Zika (2015), and severe acute respiratory syndrome coronavirus 2 (SARS-CoV-2). Five of these outbreaks were in the last decade, four of which resulted in pandemics (David M Morens and Anthony S Fauci, 2020). During viral outbreaks, there is an urgent need for diagnostic and prognostic tests for accurately diagnosing patients at high risk of severe outcome who should be admitted to hospitals, and those with mild infection who can recover at home. The ongoing SARS-CoV-2 pandemic, where approximately 80% of infected patients have mild infection, and 20% have severe illness requiring hospitalization and critical care (Wu and McGoogan, 2020), has acutely demonstrated the need for such a test to reduce the risk of hospital overrun, shortage of supplies, and the resulting socioeconomic costs.

The current armamentarium for identifying high-risk patients is comprised of lab tests (e.g., white blood cell count differentials, Procalcitonin, interleukin-6 and -8, lactate dehydrogenase, C-reactive protein) and standardized severity of illness scores designed for predicting mortality among the critically ill (e.g., PRISM, SOFA, APACHE). In a triage setting during viral outbreaks, these have limited clinical utility as they are non-specific markers of inflammation and late predictors of mortality (Falcão et al., 2019; Liu et al., 2020; Rast et al., 2014).

Several recent studies have repeatedly demonstrated the utility of the host immune response to pathogens to accurately diagnose the presence, type, and severity of infections (Andres-Terre et al., 2015; Sweeney et al., 2018a; 2015; 2016b). By leveraging clinical, biological, and technical heterogeneity across multiple independent datasets, we have previously identified a conserved generalizable host response to distinguish bacterial and viral infections (Andres-Terre et al., 2015; Sweeney et al., 2015; 2016b). We have also demonstrated that the conserved host immune response to infection can be an accurate prognostic marker of the risk of 30-day mortality in patients with infectious diseases (Sweeney et al., 2018b). These studies have further demonstrated that the host response to pathogens is detected earlier than symptom onset (Andres-Terre et al., 2015; Sweeney et al., 2016a; Warsinske et al., 2018). For example, a 3-gene host response-based signature has been repeatedly demonstrated to predict progression from latent Mycobacterium tuberculosis (Mtb) infection to active tuberculosis 6 months prior (Gupta et al., 2020; Turner et al., 2020; Warsinske et al., 2018). Finally, and most importantly, these generalizable robust host response-based signatures can be translated onto a targeted platform as a point-of-care test with rapid turnaround time (Sodersten et al., 2020).

We have previously described a conserved host response to respiratory viral infections, called the Meta-Virus Signature (MVS), that is distinct from the response to bacterial infections (Andres-Terre et al., 2015). Here, we hypothesized that the MVS is also conserved in viral infections that cause severe disease, including Ebola, SARS-CoV-2, and others. We further hypothesized that the MVS is correlated with the severity of viral infection and could be used to better distinguish patients with mild versus severe infection, irrespective of the virus. We tested these hypotheses by integrating 34 independent cohorts comprised of 4,780 blood transcriptome profiles from healthy controls and patients with acute viral infection, and single-cell RNA-seq profiles of more than 264,000 immune cells from 71 samples. We found that the conserved host response to viral infection, represented by the MVS, is (1) present in SARS-CoV-2, Ebola, chikungunya, influenza and other viruses, (2) correlated with severity of viral infection, and (3) predominantly expressed in myeloid cells. We also found the proportions of CD14+ monocytes increased and CD16+ monocytes decreased with increasing severity of viral infection across all viruses. Next, we developed a patient trajectory differentiation method using bulk transcriptome data, which found that patients with mild viral infection follow a different trajectory than those with severe viral infection. We found 96 genes significantly different between the two trajectories. These genes showed that with increased severity of viral infection, proportions of lymphoid cells (NK and T cells) decreased, whereas proportions of hematopoietic stem and progenitor cells (HSPCs) and myeloid-derived suppressor cells (MDSCs) increased, irrespective of the infecting virus. Finally, we identified four gene modules corresponding to protective and detrimental host immune responses, where a protective module was decoupled from the interferon (IFN) response in patients with severe viral infection but not in those with mild infection. Using these modules, we defined the Severe-or-Mild (SoM) score that accurately distinguished patients with non-severe and severe outcomes. Overall, by leveraging the biological, clinical, and technical heterogeneity across data, we provide strong evidence of a conserved host immune response to acute viral infection, irrespective of the virus. This conserved response presents novel opportunities to develop robust generalizable diagnostic and prognostic tests across a broad spectrum of viruses. Such a test could facilitate the risk-stratification of patients during the current pandemic and in novel outbreaks that will invariably arise in the future. Further, the coordinated immune changes observed in patients with severe viral infections in our analysis have the potential to identify novel host-directed drug targets, and present opportunities for drug repurposing.

## Results

### Data collection, curation, and preprocessing

We searched the public repositories Gene Expression Omnibus (GEO), ArrayExpress, European Nucleotide Archive (ENA), and Sequence Read Archive (SRA) for transcriptome profiles of peripheral blood samples from patients with viral infection. We excluded all datasets used to discover the MVS previously to ensure all cohorts analyzed in the current study were independent. We identified 34 independent cohorts within 26 datasets composed of 4,780 samples from patients across 18 countries infected with at least one of 16 viruses (**Figure 1A, Table S1** and **Study Summaries**). We defined a cohort as a comparable group of individuals within a dataset, where each dataset has a unique GEO identifier and may contain multiple cohorts. For example, the dataset GSE73072 contains seven cohorts of individuals challenged with one of three viruses. Overall, these cohorts included a broad spectrum of biological, clinical, and technical heterogeneity represented by blood samples profiled from children and adults infected with a virus using either microarray or RNA sequencing (**Methods**).

**Figure 1:**
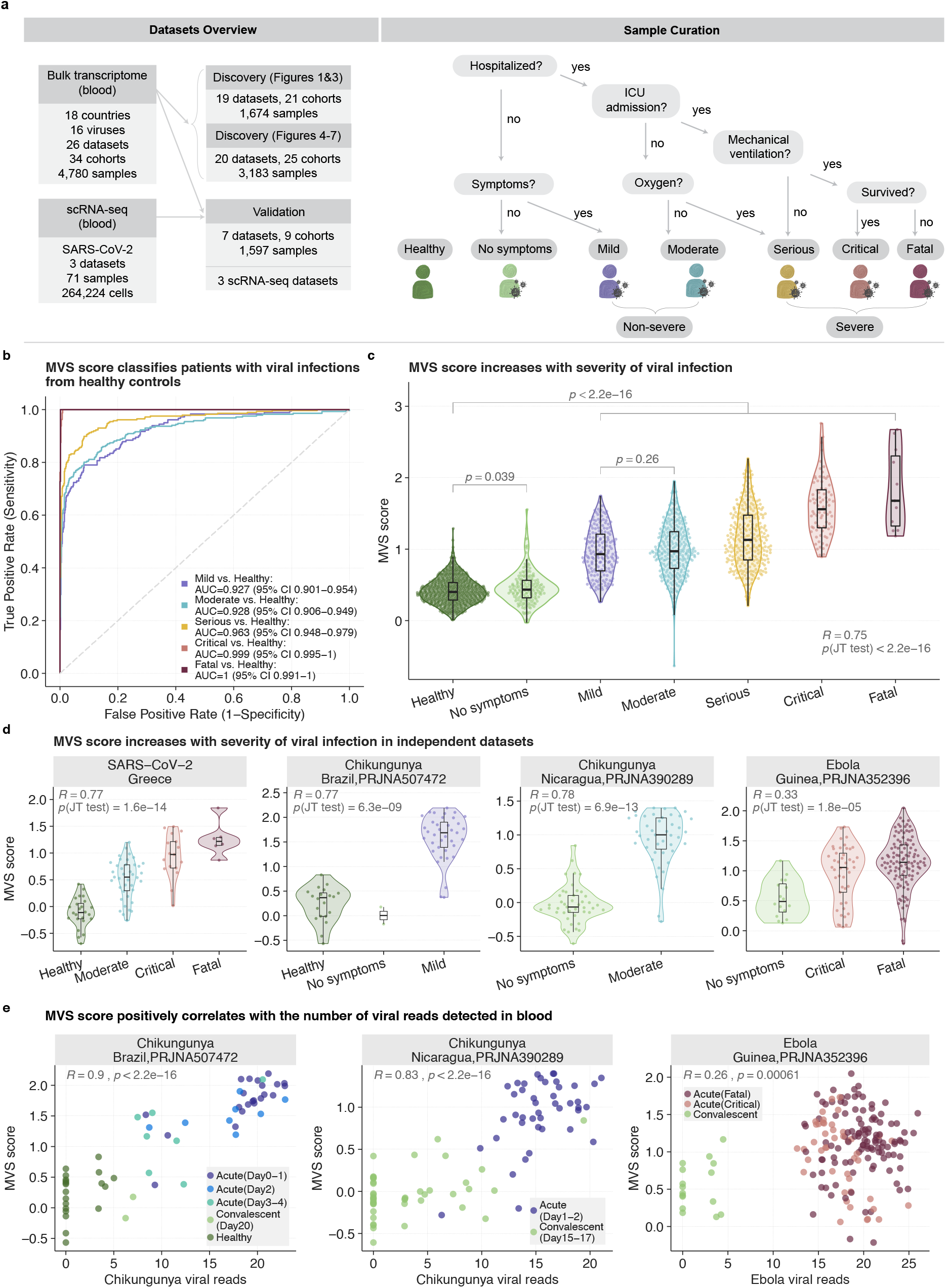
Conserved host response to viral infection, represented by the MVS, is associated with severity. **a**) Overview of datasets used for analysis (left), and criteria for assigning viral infection severity categories to samples (right). The 4,780 bulk transcriptome samples from 26 datasets were divided into discovery and validation cohorts. The “No symptoms” category includes both individuals with asymptomatic viral infection or convalescents. **b)** ROC curves for distinguishing patients with viral infection of varying severity from healthy controls using the MVS score in 1,674 samples across 19 datasets. The AUROC increased with severity of viral infection. **c**) Distribution of the MVS scores across the severity of viral infection in 1,674 samples in 19 datasets. Each point in the violin plot represents a blood sample. Jonckheere-Terpstra (JT) trend test was used to assess the significance of the trend of the MVS score over severity of infection. P-values for the comparison of MVS scores in two groups were computed using Mann–Whitney U test.**d**) Validation of correlation between the MVS score severity of viral infection in 4 independent RNA-seq datasets from patients with SARS-CoV-2, chikungunya, or Ebola infection. None of these viruses were used in the discovery, demonstrating generalizability of the MVS to previously unseen viruses. **e**) Positive correlation between the MVS score and the number of viral reads detected in blood samples in 3 independent RNA-seq datasets. Each point represents a sample. The X axis represents the number of viral reads, and the Y axis represents the MVS score for each sample.

We assigned a standardized severity category to each of the 4,780 samples (**Figure 1A** and **Methods**). Briefly, we assigned samples from individuals with no viral infection and no other disease to the “healthy” category. We assigned samples from asymptomatic individuals with confirmed acute or convalescent viral infection to the “no symptoms” category. Next, we divided symptomatic patients with viral infection into those who were hospitalized and those who were not. Patients with viral infection that were not hospitalized and managed as outpatients were categorized as “mild.” We further divided the hospitalized patients with viral infection based on whether they were admitted to the intensive care unit (ICU) or not. We assigned hospitalized patients admitted to the general wards without requiring supplemental oxygen to the “moderate” category; those on the general wards who required oxygen or those admitted to the ICU without mechanical ventilation or inotropic support were assigned to the “serious” category, and those admitted to the ICU with mechanical ventilation or inotropic support were assigned to the “critical” category. Patients who died were assigned to the “fatal” category. For cohorts that lacked sample-level severity data, we assigned the same severity category to each sample based on the cohort description. Finally, we defined two additional broader categories: “non-severe,” which included patients with mild and moderate viral infection, and “severe,” which included patients with serious, critical, and fatal viral infection (**Figure 1A**).

### MVS represents a conserved host response to viral infection and is associated with severity

To test our hypothesis that a conserved host response to viral infection, represented by the MVS score, is associated with severity, we co-normalized transcriptome profiles of 1674 blood samples (663 healthy, 167 asymptomatic or convalescent, 181 mild, 286 moderate, 286 serious, 80 critical, and 11 fatal) from 21 cohorts across 19 independent datasets using COCONUT, which removes inter–dataset batch effects while remaining unbiased to the diagnosis of the diseased patients (**Figure 1A, Table S1** and **Methods**) (Sweeney et al., 2016b).

The majority of patients in these 19 datasets were infected with adenovirus, influenza, human rhinovirus (HRV), or respiratory syncytial virus (RSV). The MVS score accurately distinguished patients with viral infection from healthy controls across all datasets as well as in individual datasets (**Figure 1B** and **Figure S1A**). The area under the receiver operating characteristics (AUROC) curves increased with severity (0.925<AUROC≤1), further suggesting that a conserved host response is associated with the severity of viral infection.

Therefore, we correlated the MVS score with standardized severity categories and found a significant correlation between the MVS score and the severity of viral infection (r=0.75, p<2.2e-16; **Figure 1C**). The MVS score was significantly higher in all infected patients compared to healthy controls (p<2.2e-16), irrespective of symptoms, severity, and virus (**Figure 1C**). In asymptomatically infected or convalescent patients, the MVS score was marginally higher than in healthy controls (p=0.039). The MVS score was not statistically different between patients with mild versus moderate severity (p=0.26). Importantly, across all datasets except one, the MVS score was significantly correlated with viral infection severity (0.43≤R≤0.93; p<0.02), irrespective of virus, geography, or age (**Figure S1B**). Furthermore, in 405 samples from patients infected with one of three viruses (SARS-CoV-2, Ebola, chikungunya) across 4 datasets that were not COCONUT co-normalized, the MVS score was correlated with severity (0.33≤R≤0.78; P≤1.8e-05; **Figure 1D**) and accurately distinguished patients with viral infection from healthy controls in each dataset (0.84≤R≤0.972; **Figure S1C**). In three independent datasets of blood samples from patients with either chikungunya or Ebola infection, profiled using RNA-seq, we detected sequencing reads from the corresponding viral RNA (**Methods**). In each of these three datasets, the MVS score significantly correlated with the number of viral reads detected in blood (p≤6.1e-4; **Figure 1E**). Further, in each dataset, both the number of viral reads in blood and the MVS score decreased as patients progressed from acute infection to convalescence.

Collectively, our results demonstrate that a conserved host response to viral infection, represented by the MVS score, is correlated with the severity of viral infection and the number of viral reads detected in blood samples from patients, irrespective of clinical, biological, or technical heterogeneity or the infecting virus.

### Myeloid cells are the primary source of MVS that correlate with the severity of viral infection

Next, to gain a mechanistic understanding of the conserved host response to viral infection, we investigated whether the MVS score is associated with specific immune cell types. We integrated three single-cell RNA-seq (scRNA-seq) datasets consisting of 264,224 immune cells from 71 PBMC samples (50 SARS-CoV-2, 17 healthy, 2 influenza, 2 RSV) from 54 individuals across three independent cohorts (Seattle, Atlanta, Stanford) (**Table S2**) (Arunachalam et al., 2020; Su et al., 2020; Wilk et al., 2020). The Seattle Cohort profiled 135,420 immune cells from 39 PBMC samples of healthy controls and patients with SARS-CoV-2 infection (6 healthy, 1 mild, 8 moderate, 17 serious, 7 critical) using CITE-seq. The patients with SARS-CoV-2 infection in the Seattle Cohort were profiled at two time points: (1) near the time of a positive clinical diagnosis and (2) a few days later. The Atlanta Cohort profiled 84,083 immune cells from 18 PBMC samples of healthy controls and patients infected with one of 3 viruses (5 healthy, 1 moderate influenza, 1 serious influenza, 2 serious RSV, 2 convalescent SARS-CoV-2, 3 moderate SARS-CoV-2, 3 serious SARS-CoV-2, 1 fatal SARS-CoV-2) using CITE-seq. Finally, the Stanford Cohort profiled 44,721 immune cells from 14 PBMC samples of healthy controls and patients with SARS-CoV-2 infection (6 healthy, 1 moderate, 3 serious, 3 critical, 1 fatal) using Seq-Well. Collectively, these three cohorts included clinical, biological, and technical heterogeneity at a single-cell level.

We integrated the three scRNA-seq cohorts using Seurat (Satija et al., 2015), and visualized the data in a low dimensional space using Uniform Manifold Approximation and Projection (UMAP) (**Figure 2A-D**). Immune cells across the three cohorts clustered into myeloid cells (monocytes, myeloid dendritic cells, granulocytes, etc.), T and NK cells, and B cells (**Figure 2A-B**). The MVS score was substantially higher in myeloid cells from hospitalized patients with viral infection (**Figure 2C-E** and **Figure S2A**) and positively correlated with the severity of viral infection in myeloid cells at the single-cell level (R=0.63, p=2e-08), which was driven by CD14+ monocytes (R=0.63, p=4.1e-08) compared to CD16+ monocytes (R=0.18, p=2.5e-03) (**Figure 2F**). Further, proportions of myeloid cells increased with severity (R=0.33, p=4.5e-03), which was also driven by increased proportions of CD14+ monocytes (R=0.47, p=1e-04). In contrast, proportions of CD16+ monocytes decreased with increasing severity of viral infection (R= -0.55, p=4.5e-06) (**Figure 2G**). Finally, the MVS score in myeloid cells at the single-cell level and proportions of myeloid cells were positively correlated (R=0.3, p=0.0097), which was also driven by CD14+ monocytes (R=0.51, p=6.7e-06) but not CD16+ monocytes (**Figure 2H**). Collectively, these results demonstrated that in response to a viral infection, proportions of myeloid cells, primarily CD14+ monocytes, increase with severity along with the conserved host response to viral infection represented by the MVS at a single-cell level.

**Figure 2:**
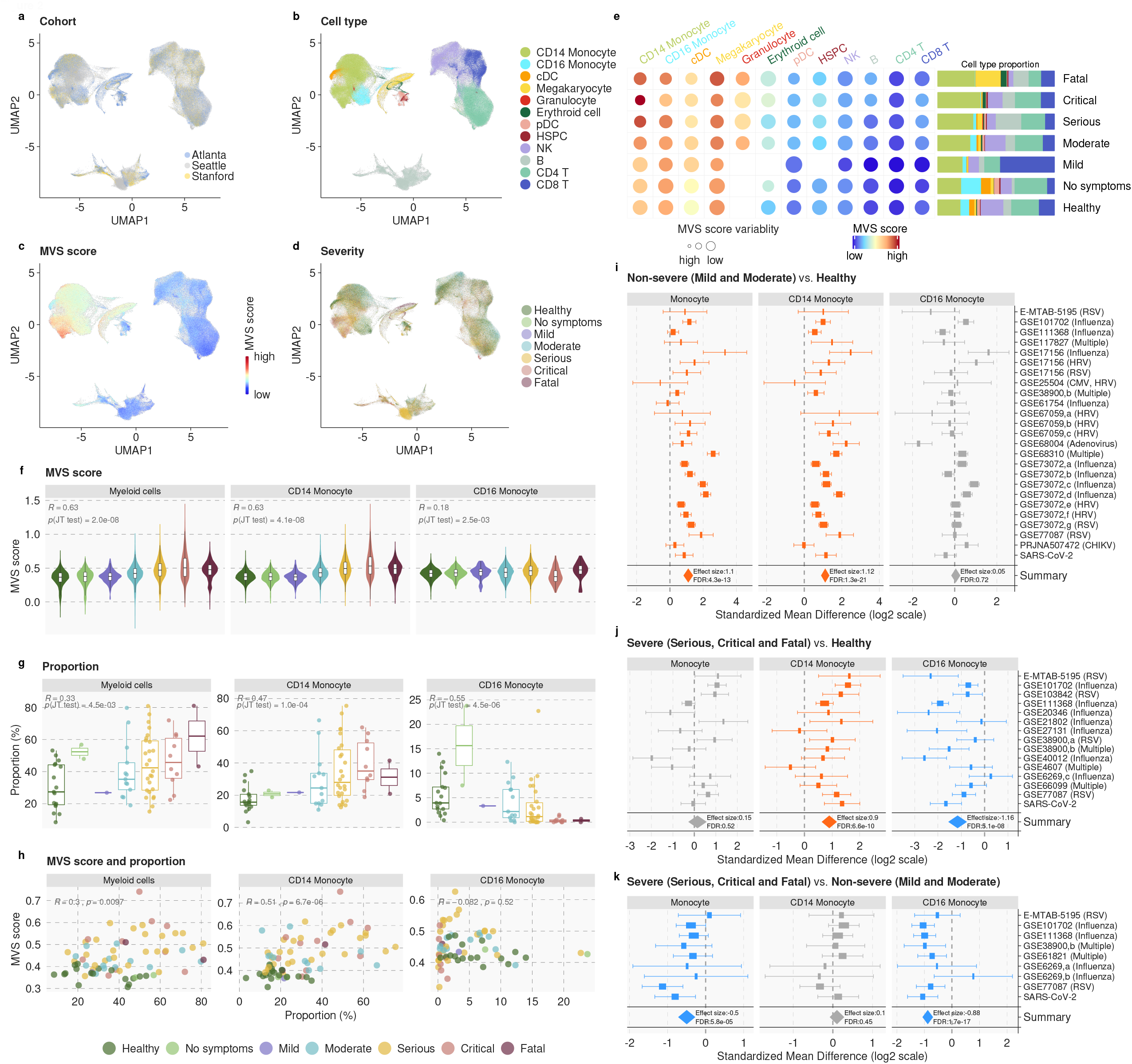
Single-cell RNA-seq identifies monocytes as the primary source of the MVS. **a-d**) The MVS score was higher in myeloid cells from hospitalized patients with viral infection (i.e., moderate, serious, critical, and fatal). UMAP visualization of 264,224 immune cells from 71 PBMC samples in three independent scRNA-seq datasets colored by **a)** cohort, **b)** cell type, **c)** MVS score, and **d)** severity of viral infection. **e**) Circle map depicting the average MVS score in each cell type in each category of viral infection severity. The color of the circle represents the average MVS score in each cell type. The size of the circle describes the variability of MVS score in the cell type, with larger size representing lower variability. The barplot on the right of the circle map shows the mean proportion of each cell type in each severity category. A detailed sample-level MVS score distribution across cell types can be found in Figure S2A. **f-h)** Proportions of myeloid cells and the MVS score at a single cell level increase with severity of viral infection predominantly driven by CD14+ monocytes. **f)** MVS score in each cell in myeloid cells, CD14+ monocytes, and CD16+ monocytes, **g)** Proportion of myeloid cells, CD14+ monocytes, and CD16+ monocytes in each individual, and **h)** correlation between mean the MVS score and cellular proportion in each individual. **i-k**) *In silico* deconvolution of bulk transcriptome profiles from healthy controls and patients with viral infection validates increased CD14+ monocytes and reduced CD16+ monocytes with increased severity in 2,027 blood samples from 32 independent cohorts. Forest plots for change in proportions of monocytes, CD14+ monocytes, and CD16+ monocytes in **i)** non-severe patients compared to healthy controls, **j)** severe patients compared to healthy controls, and **k)** severe patients compared to non-severe patients. The x axes represent standardized mean difference between two groups, computed as Hedges’ *g*, in log2 scale. The size of the rectangles is proportional to the standard error of mean difference in the study. Whiskers represent the 95% confidence interval. The diamonds represent overall, combined mean difference for a given cell type in a given comparison. Width of the diamonds represents the 95% confidence interval of overall mean difference.

Next, we investigated whether these changes in CD14+ and CD16+ monocytes were also observed in patients with viral infection at the bulk transcriptome level. We performed *in silico* cellular deconvolution of blood transcriptome profiles of 4357 patient samples from 32 independent cohorts using immunoStates (Bongen et al., 2018; Chowdhury et al., 2018; Scott et al., 2019; Vallania et al., 2018) (**Table S3**) to estimate proportions of 25 immune cell subsets in each sample. Then, we performed three multi-cohort analyses to compare changes in immune cell proportions in (1) non-severe viral infections compared to healthy controls, (2) severe viral infections compared to healthy controls, and (3) severe compared to non-severe viral infections.

In line with our scRNA-seq analysis, proportions of total monocytes were significantly higher in patients with non-severe viral infection compared to healthy controls (ES=1.10, FDR=4.33e-13), but were not significantly higher in those with severe viral infection (**Figure 2I-J, Table S4**). Further analysis of monocytes found the proportion of CD14+ monocytes increased significantly in patients with non-severe (ES=1.12, FDR=1.30e-21) and severe (ES=0.9, FDR=6.56e-10) viral infection compared to healthy controls, but were not significantly different between patients with non-severe and severe viral infection (**Figure 2I-K, Table S4**). In contrast, and in line with the scRNA-seq analysis, proportions of CD16+ monocytes were significantly lower in patients with severe viral infection compared to healthy controls (ES = -1.16, FDR=5.13e-08) and those with non-severe viral infection (ES= -0.88, FDR=1.73e-17) (**Figure 2J-K, Table S4**), but were unchanged in non-severe patients compared to healthy controls (**Figure 2I, Table S4**).

Cellular deconvolution analysis also found that the proportions of neutrophils were significantly higher in patients with severe viral infection compared to healthy controls (ES=1.24, FDR=4.12e-16) and those with non-severe viral infection (ES=0.99, FDR=4.33e-07) (**Figure S2B, Table S4**). These results were in line with the scRNA-seq data, where we detected granulocytes only in patients with moderate through fatal viral infection (**Figure 2E, Figure S2A**). These cells are likely low-density immature granulocytes typically found in patients with sepsis. These results also demonstrated the complementarity of *in silico* deconvolution to scRNA-seq, which does not allow the profiling of neutrophils.

Collectively, our integrated analyses of scRNA-seq from 3 cohorts of 71 PBMC samples and *in silico* cellular deconvolution of bulk transcriptome profiles from 4357 samples across 32 independent cohorts using immunoStates showed that the conserved host response to viral infections is predominantly from myeloid immune cells. We also found that proportions of CD14+ monocytes increased and CD16+ monocytes decreased with increased severity of viral infection.

### MVS identifies distinct clusters of patients with non-severe and severe viral infection

Despite the consistently significant correlation between the MVS score and severity of viral infection, there was a substantial overlap in the MVS score between patients with non-severe and severe viral infection (**Figure 1C**). However, low dimensional visualization of 1674 co-normalized samples using UMAP showed that healthy controls were distinct from patients with viral infection irrespective of the infecting virus (**Figure 3A**). Importantly, the MVS score increased along the first UMAP component (UMAP1; **Figure 3B**), while patients with mild viral infection were clustered separately from those with severe viral infection along the second UMAP component (UMAP2; **Figure 3C**). We further validated the robustness of the clusters observed in UMAP by mapping 8 independent cohorts consisting of 2,604 samples from patients with one of 4 viral infections to the same low dimensional space (**Figure 3D and 3E**). All but one of these 8 cohorts were challenge studies, where 129 healthy individuals were inoculated with either influenza (1,465 samples from 70 subjects), RSV (419 samples from 20 subjects), or HRV (634 samples from 39 subjects). Each of the infected subjects in these challenge studies had asymptomatic or mild infection. When mapped to the UMAP space created using the 1,674 samples, samples from the challenge studies clustered with mild viral infections (**Figure 3D**). We also mapped 86 of the samples (24 healthy controls, 62 SARS-CoV-2), which were profiled using RNA-seq and included patients with moderate, critical, and fatal SARS-CoV-2 infection (**Figure 3E**). Patients with critical and fatal SARS-CoV-2 infection mapped to the region enriched for patients with critical and fatal viral infection, whereas patients with moderate SARS-CoV-2 infection mapped to the region enriched for patients with non-severe viral infection (**Figure 3E**), again demonstrating that the host response to viral infection is conserved and associated with severity, irrespective of the virus. Collectively, this observation suggested that a distinct subset of genes in the MVS may be differentially associated with the severity of viral infection.

**Figure 3:**
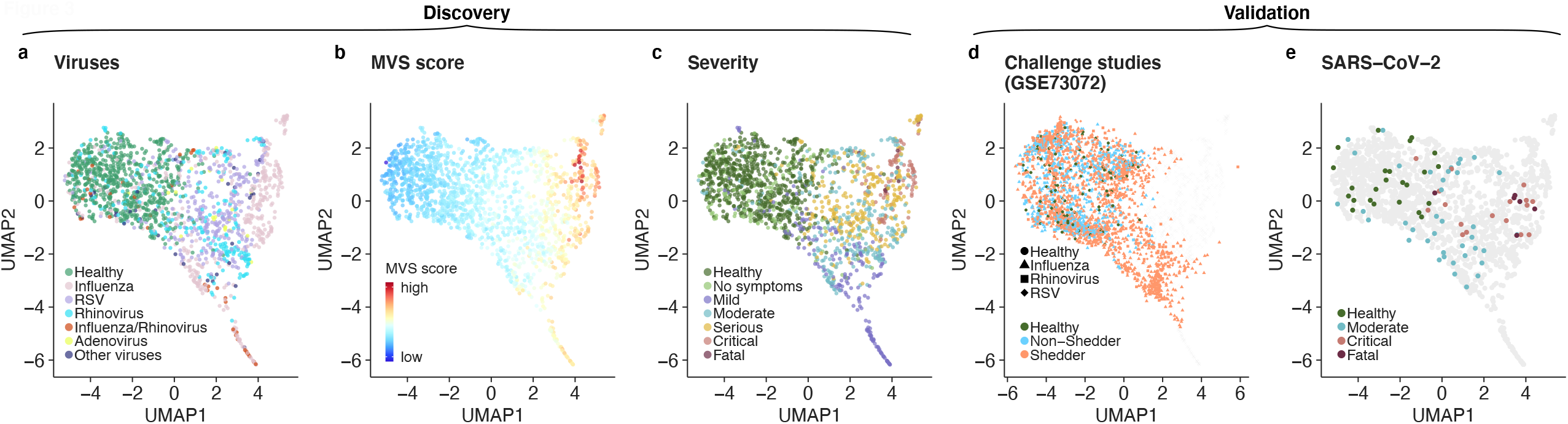
The MVS identifies distinct clusters of patients with non-severe and severe viral infection. **a-c**) UMAP visualizations of 1,674 samples in 19 datasets colored by **a)** virus **b)** MVS score, and **c)** severity of viral infection show distinct clusters of healthy controls and patients with non-severe and severe viral infection. **d-e**) Projection of independent cohorts on the UMAP space obtained from the discovery cohort: **d)** 2,518 samples from seven challenge studies using influenza, RSV, or HRV in GSE73072 and **e)** 86 samples from the SARS-CoV-2 cohort.

### Hospitalized patients with viral infection follow a different trajectory from non-hospitalized patients with viral infection

Based on low dimensional mapping of samples, we hypothesized that patients with mild and severe viral infection follow different trajectories. Each sample in our analysis represents a snapshot of the host response to viral infection that spans from recognizing the presence of a virus to its elimination. This is analogous to cellular differentiation analysis using single-cell profiling data, where each cell represents a snapshot along the differentiation trajectory. Therefore, we tested our hypothesis by adapting tSpace (Dermadi et al., 2020), a method for identifying cellular differentiation trajectories using single-cell data, to identify disease trajectories using bulk RNA data. We refer to the modified method as ‘disease space’ (dSpace) (**Methods**).

Because the viral challenge cohorts included a large number of longitudinal samples that can aid in a more accurate inference of the host response trajectories, we randomly selected four of the seven challenge studies (1,509 samples across 2 influenza, 1 HRV, and 1 RSV studies) and co-normalized them with 1674 samples from 19 datasets using COCONUT. Overall, we applied dSpace to 3183 COCONUT co-normalized samples from 25 independent cohorts. We did not include all challenge studies when inferring the disease trajectories to avoid introducing class imbalance because subjects in the challenge studies only had mild viral infections. We used these left-out challenge studies for validation of the inferred trajectories.

The first principal component of dSpace (dPC1) correlated with the severity of viral infection, whereas the second component (dPC2) distinguished hospitalized patients with viral infection from non-hospitalized patients with mild infection (**Figure 4A**). Importantly, participants from the influenza, RSV, and HRV challenge studies clustered almost exclusively with patients with mild infection (**Figure S3A**).

**Figure 4:**
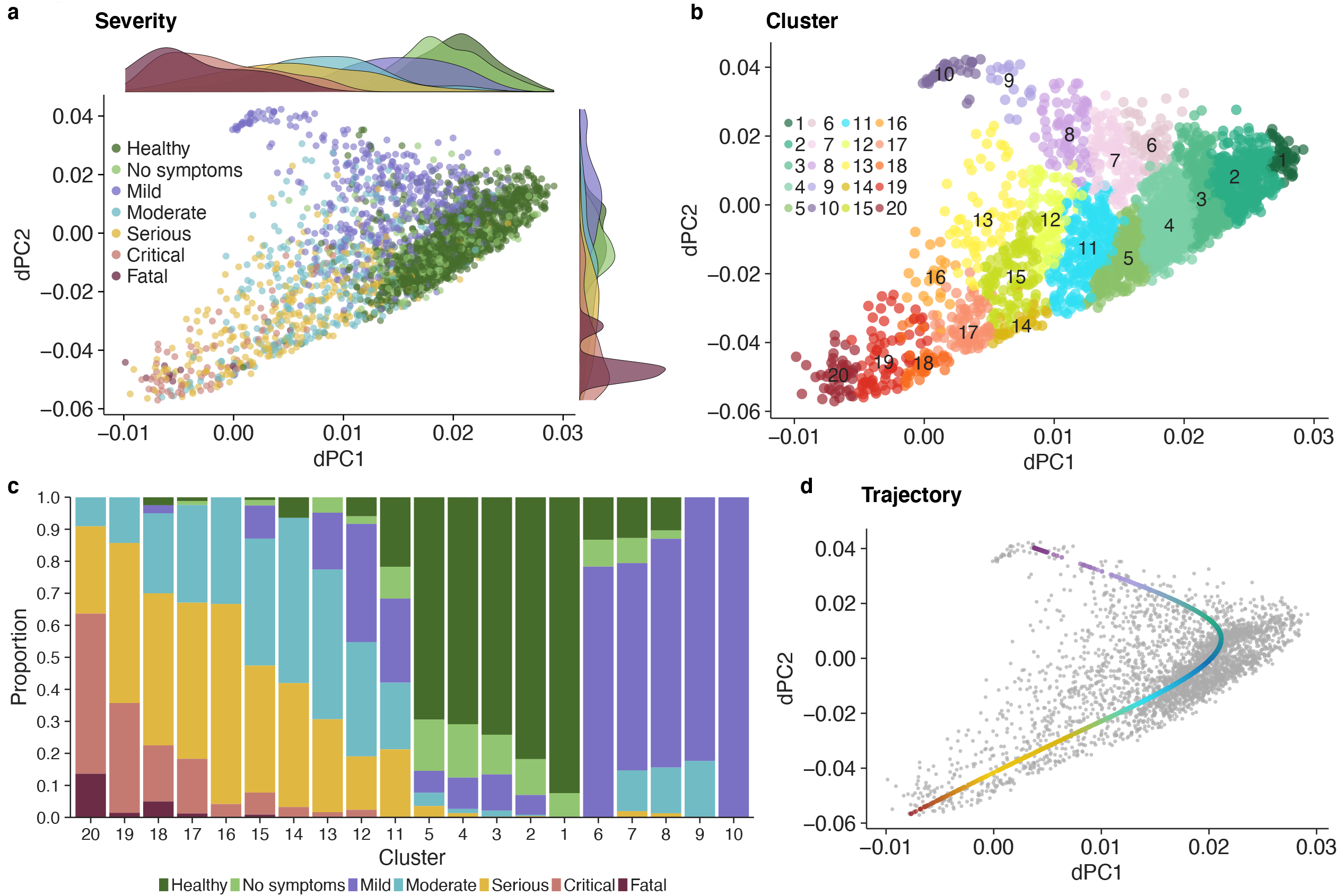
Patients with non-severe and severe viral infection follow divergent disease trajectories. **a**) Trajectory analysis of 3,183 samples from 25 cohorts using dSpace. The first two principal components of dSpace distinguish the samples by severity category. **b)** Clustering of samples using dSpace. **c)** Proportion of samples for each severity category in each cluster. Clusters 1-5 were predominantly composed of healthy controls and patients with asymptomatic viral infection or convalescents, clusters 6-10 and 13-20 were predominantly composed of patients with non-severe and severe viral infection. **d)** A principal line on dSpace coordinates identified by trajectory analysis. The red and purple colors of the line ends indicate the severe and non-severe trajectories, respectively.

Next, we clustered samples using the disease space matrix, and used the resulting clusters to isolate trajectories associated with the severity of viral infection (**Methods**). We identified 20 clusters that identified 3 groups such that one category of samples dominated (**Figure 4B**): clusters 1-5, in which healthy controls and asymptomatically infected or convalescent patients accounted for >80% of samples; clusters 6-10, in which patients with mild viral infection accounted for >68% of samples, and clusters 13-20, in which hospitalized patients with moderate, serious, critical, or fatal viral infection accounted for >77% of samples (**Figure 4C**). Clusters 11 and 12 were heterogeneous as no one group of samples dominated them. Strikingly, 1507 out of 1509 samples (99.9%) from the influenza, RSV, and HRV challenge studies fell within clusters 1-12 (**Figure S3B**), demonstrating the robustness of the clusters defined using dSpace. We fit a principal trajectory line to the dSpace matrix, which consisted of healthy patients in the center and two divergent trajectories: one dominated by patients with mild viral infection and the other dominated by hospitalized patients with viral infection (**Figure 4D, Methods**). Hitherto, we refer to these trajectories as “mild trajectory” and “severe trajectory,” respectively. Collectively, trajectory analysis using dSpace demonstrated that hospitalized patients with viral infection follow a different trajectory than those with mild infection compared to healthy controls, irrespective of the infecting virus.

### Proportions of NK cells and the expression of NK cell-specific genes are negatively correlated with the severity of viral infection

We identified 96 genes within the MVS that were significantly different between the two trajectories (**Figure 5A**). We obtained expression of each of the 96 genes in a cell type, relative to all other cell types, from the MetaSignature database (https://metasignature.stanford.edu) (**Figure 5B**) (Haynes et al., 2017; Vallania et al., 2018). The majority of the genes negatively correlated with the severity of viral infection are preferentially expressed in lymphocytes (T cells, B cells, and NK cells), whereas the majority of the genes positively correlated with severity are preferentially expressed in myeloid cells (granulocytes, monocytes, mDCs, and macrophages).

**Figure 5:**
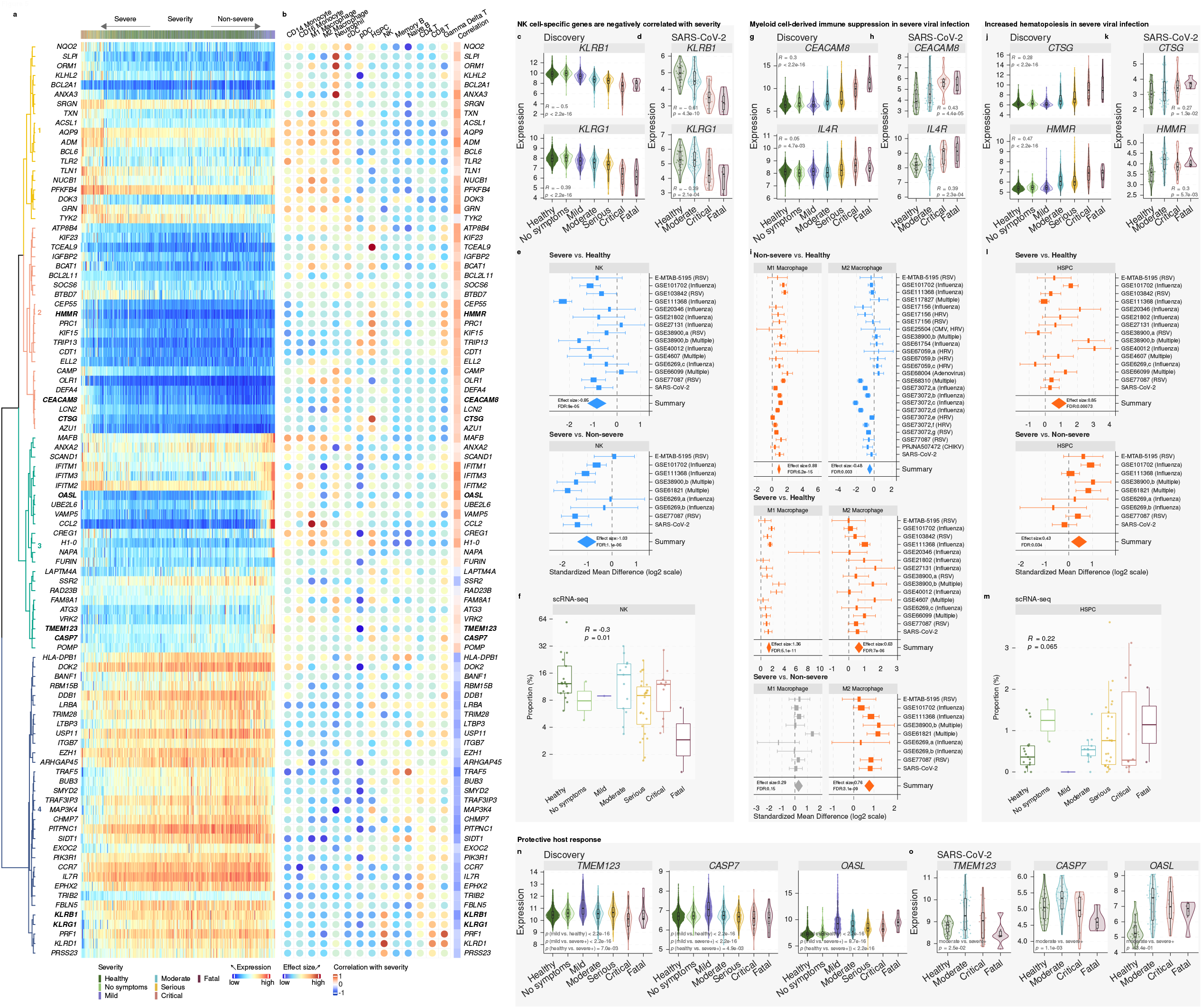
Immune responses from NK cells, myeloid cell-derived suppression, and hematopoiesis are associated with severity of viral infection. Violin plots: each dot represents a sample, and the Y-axis represents expression of the corresponding gene in a sample. Box plots: each dot represents a sample, and the Y-axis represents proportion of the corresponding cell type in a sample. Forest plots: represent comparison of change in proportions between two groups for a given immune cell type, obtained by *in silico* cellular deconvolution of blood samples, where the X-axis represents standardized mean difference between two groups, computed as Hedges’ *g*, in log2 scale. The size of the rectangles is proportional to the standard error of mean difference in the study. Whiskers represent the 95% confidence interval. The diamonds represent overall, combined mean difference for a given cell type in a given comparison. Width of the diamonds represents the 95% confidence interval of overall mean difference. **a)** Gene expression heatmap of the 96 severity trajectory-defining genes. Rows represent genes and columns represent samples, ordered by position along the disease trajectory. The dendrogram represents hierarchical clustering performed on the rows of the heatmap. Colors of the dendrograms indicate clusters of genes based on the relationship between each gene expression profile and severity of infection. **b)** Effect size of each gene, computed as Hedge’s *g*, in a given cell type compared to all other cell types and correlation of each gene with severity of viral infection. **c-d)** Expression of NK cell-specific genes is negatively correlated with the severity of viral infection in **c)** 3,183 samples across 25 cohorts used for discovery and **d)** an independent cohort of 86 samples from healthy controls and patients with SARS-CoV-2 infection used for validation. **e)** Proportions of NK cells were significantly lower in patients with severe viral infection compared to healthy controls (top panel) and non-severe viral infection (bottom panel). **f)** Proportions of NK cells reduce with severity of viral infection in three independent scRNA-seq cohorts. **g-i)** Proportions of MDSCs are higher in patients with severe viral infection. **g-h)** Expression of *CEACAM8*, a marker of PMN-derived MDSCs, and *IL4R*, a marker of monocyte-derived MDSC, increases with severity of viral infection in **g)** 3,183 samples across 25 cohorts used for discovery and **h)** an independent cohort of 86 samples from healthy controls and patients with SARS-CoV-2 infection used for validation. **i)** *In silico* cellular deconvolution analysis showed proportions of pro-inflammatory macrophages (M1) increase during viral infection irrespective of severity, but proportions of anti-inflammatory macrophages (M2) decrease in patients with non-severe viral infection, and increase in patients with severe viral infection. **j-k)** Expression of HSPC-specific genes is positively correlated with the severity of viral infection in **j)** 3,183 samples across 25 cohorts used for discovery and **k)** an independent cohort of 86 samples from healthy controls and patients with SARS-CoV-2 infection used for validation. **l)** Proportions of HSPCs were significantly higher in patients with severe viral infection compared to healthy controls (top panel) and non-severe viral infection (bottom panel). **m)** Proportions of HSPCs increase with severity of viral infection in three independent scRNA-seq cohorts. **n-o)** Genes expressed at higher level in patients with mild or moderate viral infection compared to healthy controls and those with severe viral infection in **n)** 3,183 samples across 25 cohorts used for discovery and **o)** an independent cohort of 86 samples from healthy controls and patients with SARS-CoV-2 infection used for validation.

Trajectory analysis identified several NK cell-specific genes from the killer cell lectin-like receptor (KLR) family (*KLRB1, KLRG1, KLRD1*) and phosphoinositide-3-Kinase (PI3K) signaling genes (*PIK3R1*), which negatively correlated with severity (**Figure 5C** and **Figure S4A**). PI3K signaling in NK cells and mutations in *PIK3R1* have been linked with human immunodeficiency and viral infections (Mace, 2018). These genes were also significantly lower in critical and fatal SARS-CoV-2 infections compared to healthy controls (**Figure 5D and Figure S4B**). Therefore, we hypothesized that NK cell proportions decreased with increased severity of viral infection. *In silico* cellular deconvolution analysis of bulk transcriptome profiles using immunoStates showed the proportions of NK cells were significantly lower in patients with severe viral infections compared to healthy controls (ES= - 0.85, FDR=8.97e-05) and non-severe viral infections (ES= -1.03, FDR=1.13e-06) (**Figure 5E, Table S4**). Further, across the three independent cohorts profiled using scRNA-seq, NK cell proportions in severe patients were lower than in healthy controls (**Figure 5F**). Collectively, trajectory analysis using dSpace, deconvolution using immunoStates, and scRNA-seq found that the proportions of NK cells and the expression of several NK cell-associated genes reduced with increased severity of viral infection, irrespective of the infecting virus.

### Myeloid-derived immune suppression is higher in patients with severe viral infection

In line with the positive correlation between the MVS score, proportions of myeloid cells, and infection severity, several differentially expressed genes between the two trajectories (**Figure 5A**) were preferentially expressed by immune cells of the myeloid lineage (**Figure 5B**). As expected, a subset of positively correlated genes with viral infection severity (*CAMP, BCAT1, LCN2, TXN*) have known proinflammatory functions in myeloid cells (Bertini et al., 1999; Bruns et al., 2015; Choi and Fujii, 2019; Eriksson et al., 2017; Papathanassiu et al., 2017; Ramos-Martínez et al., 2018) (**Figure S4C)**.

However, we also found strong evidence of increased myeloid cell-derived immune suppression in patients with severe viral infection. First, markers of polymorphonuclear myeloid-derived suppressor cells (PMN-MDSCs), *CEACAM8* (*CD66B*; **Figure 5G**) and *OLR1* (*LOX-1;* **Figure S4C**), were higher in patients with severe viral infection. Second, markers of monocytic MDSCs (M-MDSCs), *IL-4R* (**Figure 5G**), *ITGAM* (*CD11B;* **Figure S4D**), and a functional marker of MDSCs, *ARG1* (**Figure S4D**), were also positively correlated with the severity of viral infection. Third, *ORM1*, which drives the differentiation of monocytes to anti-inflammatory M2b macrophages (Nakamura et al., 2015), was significantly different between the two trajectories. Fourth, genes known to reduce the type I interferon response, *GRN* and *BCL6* (Wei et al., 2019; Wu et al., 2016), were positively correlated with severity (**Figure S4C**). Most importantly, all genes but *GRN* positively correlated with severity in the independent cohort of patients with SARS-CoV-2 infection (**Figure 5H** and **Figure S4D**). Notably, *ORM1* expression was significantly lower in mild patients but significantly higher in severe patients compared to healthy controls (**Figure S4C**). *ORM1* expression also showed the same trends in the independent cohort of patients with SARS-CoV-2 infection, although this was not statistically significant (**Figure S4D**).

Based on these multiple lines of evidence, we hypothesized that proportions of pro- and anti-inflammatory macrophages would differ between patients with non-severe versus severe viral infection. To test this hypothesis, we extended our *in silico* cellular deconvolution analysis using immunoStates. Proportions of pro-inflammatory (M1) macrophages were higher in patients with non-severe (ES=0.88, FDR=6.16e-15) and severe (ES=1.36, FDR=5.12e-11) viral infection compared to healthy controls (**Table S4**). In contrast, when compared to healthy controls, proportions of anti-inflammatory (M2) macrophages were lower in non-severe patients (ES= - 0.48, FDR=3.00e-03), but higher in severe patients (ES=0.63, FDR=7.02e-06) (**Figure 5I, Table S4**). Proportions of M2 macrophages were also higher in severe patients compared to non-severe patients (ES=0.76, FDR=3.12e-09), but proportions of M1 macrophages were not statistically different (**Figure 5I, Table S4**). Collectively, trajectory analysis and *in silico* deconvolution provide strong evidence of increased myeloid-derived immune suppression in patients with severe viral infection. Importantly, these results suggest MDSCs as potential therapeutic targets for patients with severe viral infection.

### Increased hematopoiesis in patients with severe viral infection

Several significantly different genes between the two trajectories (*CTSG, PRC1, DEFA4, KIF15, TCEAL9, HMMR, CEP55*, and *AZU1*) were over-expressed in patients with severe viral infection, but not in those with non-severe viral infection compared to healthy controls (**Figure 5J** and **Figure S4E**). Using the MetaSignature database, we found all but one of these genes (*DEFA4*) are preferentially expressed at significantly higher levels in circulating HSPCs (**Figure 5B**). These genes were also over-expressed in patients with severe SARS-CoV-2 viral infection in an independent cohort (**Figure 5K** and **Figure S4F**). Therefore, we investigated whether HSPCs were higher in patients with severe viral infection, but not in those with non-severe viral infection. Deconvolution analysis using immunoStates found that HSPCs were significantly higher in patients with severe viral infection compared to healthy controls (ES=0.85, FDR=7.33e-04) and compared to patients with non-severe viral infection (ES=0.43, FDR=3.38e-02) (**Figure 5L, Table S4**), but not in those with non-severe viral infection compared to healthy controls (**Table S4**). Finally, in line with the trajectory and deconvolution analyses, the proportions of HSPCs increased with severity in scRNA-seq across three independent cohorts of patients with SARS-CoV-2 (**Figure 5M**).

### Trajectory analysis identifies a protective host response associated with mild viral infections

Finally, dSpace analysis identified several genes (*CCL2, OASL, CASP7, TMEM123, MAFB, VRK2, UBE2L6, NAPA*) significantly higher in patients with mild viral infection than those with severe viral infection or healthy controls (**Figure 5N** and **Figure S4G**). Specifically, we observed high expression of *CCL2*, a type I interferon receptor-mediated chemoattractant, which promotes monocyte migration to the site of infection, and *OASL*, a type I interferon-induced gene, in patients with mild viral infection. *CASP7* is cleaved by *CASP3* and *CASP10*, and is activated upon cell death stimuli and induces apoptosis. *TMEM123*, also known as *PORIMIN*, is a cell surface receptor that mediates oncosis, a type of cell death distinct from apoptosis characterized by a loss of cell membrane integrity without DNA fragmentation. These genes were also under-expressed in patients with severe SARS-CoV-2 viral infection compared to those with moderate infection (**Figure 5O** and **Figure S4H**). Collectively, these results suggest that patients with better ability to recruit monocytes, respond to interferon, and increased cell death are at a lower risk of severe viral infection.

### Coordinated protective and detrimental host response modules are associated with the severity of viral infection

Unsupervised hierarchical clustering grouped the 96 genes from dSpace analysis into four distinct modules (**Figure 5A**). Module 1 and 2 were composed of genes preferentially expressed in myeloid and HSPCs, and were higher in patients with severe viral infection (**Figure 5B)**, whereas module 4 was composed of genes preferentially expressed in lymphoid cells (NK, T, and B cells) and were higher in patients with mild viral infection compared to those with severe infection (**Figure 5B**). Module 3 included genes expressed at higher levels in patients with mild viral infection. Therefore, these four modules broadly divided the host response genes differentially expressed between two trajectories into two categories: a detrimental host response represented by module 1 and 2 (higher in patients with severe viral infection), and a protective host response represented by module 3 and 4 (higher in patients with mild viral infection).

We selected 42 out of 96 genes with significantly high effect size (|effect size|≥1) between the severe and mild trajectories (**Table S5**), which included 11, 13, 10, and 8 genes in modules 1, 2, 3, and 4, respectively. Module scores, defined as the geometric mean of expression of genes in a given module, using these reduced sets of genes continued to be significantly positively (module 1, 2, and 3) and negatively (module 4) correlated with severity of viral infection (|r| ≥ 0.43, p<2.23-16; **Figure 6A**), which further suggested that genes within each module are correlated with each other. Indeed, we found most pairs of genes within each module were positively correlated, irrespective of their infection status (**Figure 6B**).

**Figure 6:**
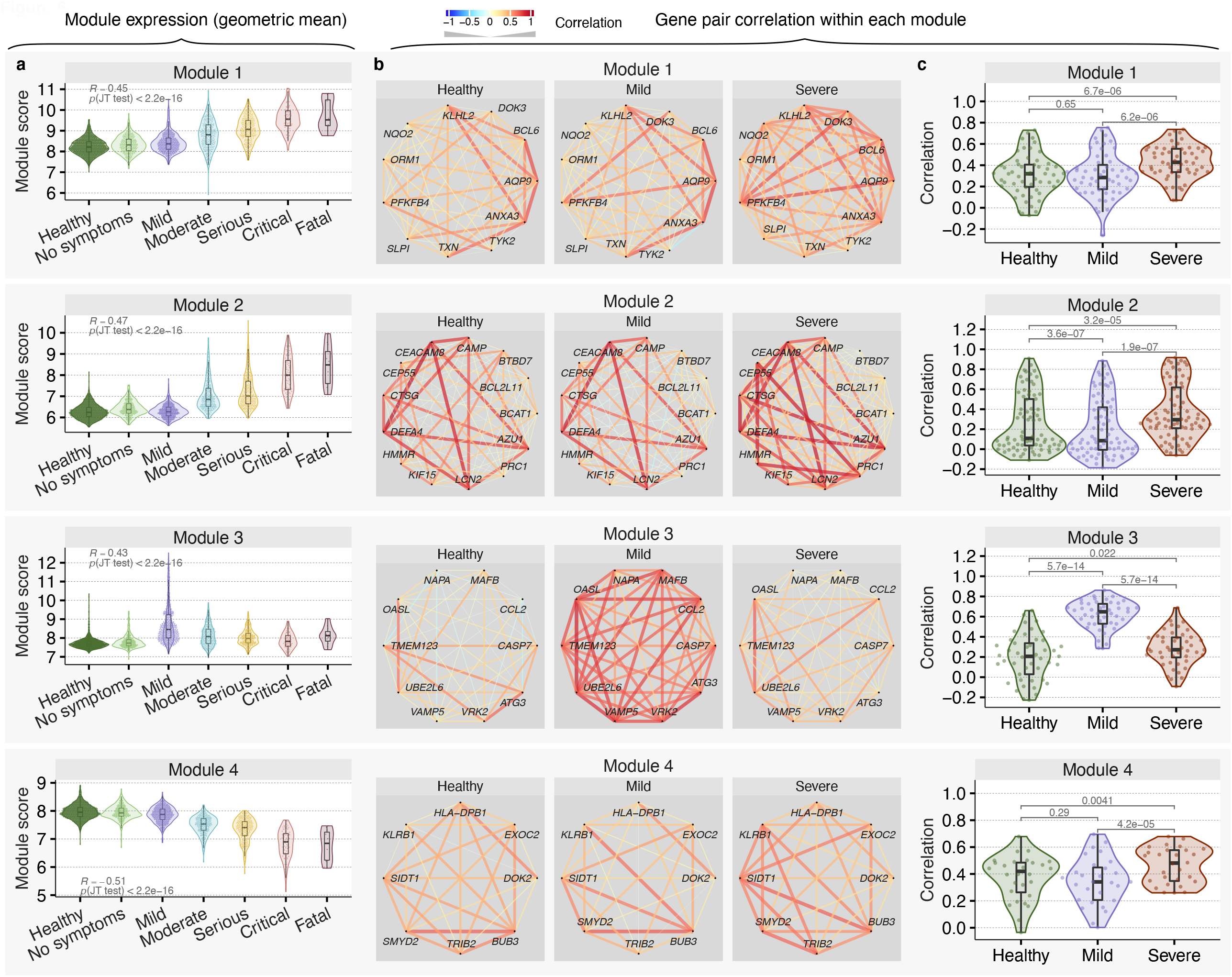
Coordinated protective and deleterious host response modules associated with severity of viral infection. **a**) The module score for 3,183 samples from 25 cohorts in the four modules. The module score of a sample is defined as the geometric mean of expression of genes in each module in the sample. **b-c**) Pairwise Spearman’s rank correlation coefficient between genes in each module in healthy controls and patients with mild or severe viral infection. **b)** The width and color of a line connecting two genes represents a correlation value between two genes. The width of the line indicates strength of correlation; red and blue color indicate positive and negative correlation, respectively. **c)** Genes in modules 1, 2 and 4 are more correlated with each other in patients with severe infection than mild infection, whereas genes in module 3 are more correlated with each other in patinents with mild infection than severe infection. Each dot in the violin plots represents the correlation between a pair of genes. P-values for the comparisons in the violin plots were computed using Wilcoxon signed-rank test.

Interestingly, we found the correlation structure within each module changed depending on the presence and severity of infection. The distribution of pairwise correlations between genes in modules 1, 2, and 4 was significantly higher in patients with severe viral infection than healthy controls or patients with mild viral infection (p<5e-05; **Figure 6C**). Interestingly, the distribution of pairwise correlations in module 2 was significantly lower in patients with mild infection compared to healthy controls (p=3.6e-07; **Figure 6C**). In contrast, pairwise correlations between genes in module 3, which included genes involved in the protective host response, were significantly higher in patients with mild infection compared to healthy controls and those with severe infection (p=5.7e-14; **Figure 6C**). Collectively, these results demonstrate that the genes within each module are expressed in a coordinated manner depending on the infection status and severity of infection.

### The protective host response module is decoupled from the interferon response in patients with severe viral infection

Recent reports have described higher expression of interferon-stimulated genes (ISGs) in patients with moderate SARS-CoV-2 infection than those with severe infection (Arunachalam et al., 2020). Therefore, we investigated whether this observation is generalizable to other viruses. Indeed, module 3 included three interferon-induced transmembrane (IFITM) genes (*IFITM1, IFITM2, IFITM3*), involved in the restriction of multiple viruses (Bailey et al., 2014), that were over-expressed in patients with viral infection and positively correlated with severity (**Figure S5A**). We also found several type I and II interferon receptors over-expressed during viral infection that positively correlated with severity, irrespective of the infecting virus (**Figure S5A**). Strikingly, in patients with mild viral infection, the distribution of correlations between IFITMs and genes in the protective response module 3 was significantly higher than in patients with severe viral infection or healthy controls (p≤1e-06; **Figure S5B**). Further, the distribution of correlations between the type I and II interferon receptors and the protective response module 3 was not statistically different between healthy controls and patients with severe viral infection, but was significantly higher in patients with mild viral infection (**Figure S5C**). These results show that while expression of the ISGs increase with the severity of viral infection, their correlation with the protective host response does not increase in patients with severe viral infection as much as those with mild viral infection. Collectively, these results demonstrate a decoupling of the protective host response from the interferon response in patients with severe viral infection, irrespective of the virus.

### Host response-based module score improves classification of patients with severe and non-severe viral infection

Despite correlating significantly with the severity of viral infection, the MVS score cannot separate severe from non-severe patients with clinically relevant accuracy (**Figure S6A-B**). We hypothesized that a score that considers the distinction between the protective and detrimental host responses would improve discrimination between patients with severe and non-severe viral infection. The two deleterious host response module scores (modules 1 and 2) were higher in samples along the severe trajectory than those along the mild trajectory, whereas the two protective host response module scores (modules 3 and 4) had the opposite pattern of expression (**Figure 7A**). We defined the Severe-or-Mild (SoM) score of a sample as the sum of the scores for module 1 and 2 divided by the sum of the scores for module 3 and 4 (**Methods**). The SoM score showed a more pronounced gradient between the severe and mild trajectories than any of the individual module scores (**Figure 7B**). Indeed, across the 3183 samples used for discovery of the trajectories, the SoM score distinguished patients with mild infection from those with severe infection with AUROC ≥ 0.929 (**Figure 7C, Figure S6C**). Importantly, the SoM score also distinguished patients with mild infection from those with severe infection with very high accuracy (AUROC>0.98) in 5 independent validation cohorts comprised of 1154 samples from patients infected with 4 different viruses (SARS-CoV-2, influenza, HRV, chikungunya) (**Figure 7D, Figure S6D**). Importantly, in patients with non-severe viral infection, the SoM score distinguished those with mild infection from those with moderate infection with higher accuracy (AUROC>0.75) compared to the MVS score (AUROC<0.63) in discovery and validation datasets (**Figure S6A-D**). Further, in hospitalized patients with a viral infection, the SoM score also distinguished those with moderate infection from those with critical or fatal infection with significantly higher accuracy than the MVS score (**Figure S6A-D**). Collectively, our results demonstrate that the protective and detrimental host response modules identified by trajectory analysis improve discrimination accuracy between patients with mild, moderate, and severe viral infection. These results further suggest that suppressing the detrimental host response modules or enhancing the protective host response modules could be therapeutic targets for host-directed broad-spectrum intervention in patients with severe viral infection.

**Figure 7:**
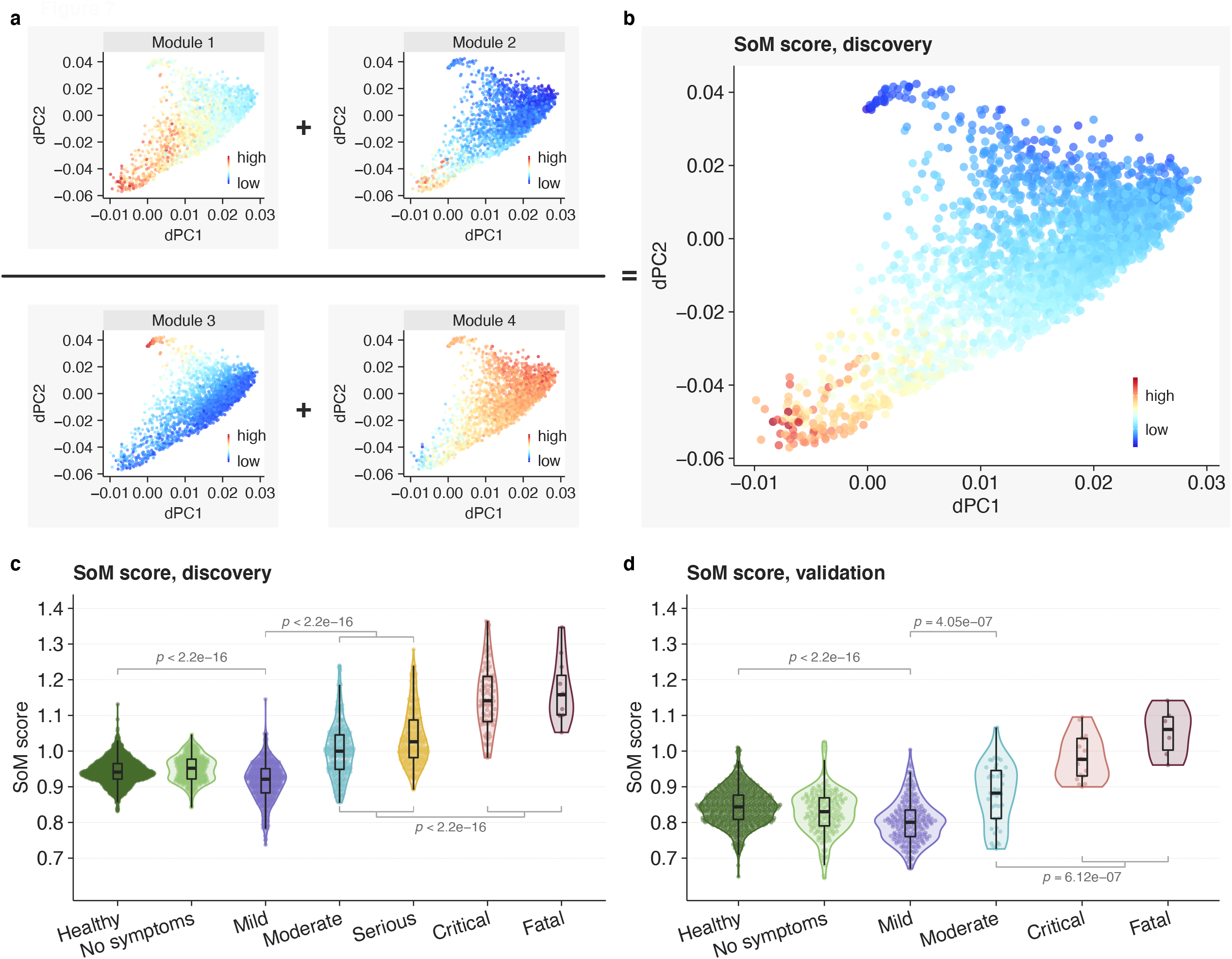
Host response modules improve classification of patients with severe and non-severe viral infection. **a)** Each of the four module scores across the 3,183 dSpace samples. **b**) The SoM score is calculated by taking the sum of the module 1 and 2 scores divided by the sum of the module 3 and 4 scores. **c-d**) The SoM score distinguishes mild and severe viral infection in the **c)** discovery cohort and **d)** validation cohort. Each point in the violin plots represents a sample. P-values for the comparisons of SoM scores between groups were computed using Mann–Whitney U test.

## Discussion

The four pandemic viral outbreaks between 2009 and 2019 have underscored an urgent unmet need for identifying generalizable diagnostic and prognostic tests. As the SARS-CoV-2 pandemic has shown, a test that could be readily deployed for triage to identify patients at high risk of severe outcome could avoid the catastrophic socioeconomic costs associated with overwhelming healthcare systems worldwide. Here, we tested a hypothesis that our previously described conserved host response to viral infections is associated with severity, and can identify patients at higher risk of severe outcome. Such a host response-based diagnostic and prognostic test could aid in the current pandemic, and allow us to be better prepared when the next viral outbreak occurs.

To test this hypothesis, we performed the largest, most comprehensive systems immunology analysis of blood transcriptome profiles from patients with viral infection to date by integrating 4,780 blood transcriptome profiles from patients with one of 16 viral infections across 34 independent cohorts from 18 countries. Further, we integrated scRNA-seq profiles of more than 264,000 immune cells from 71 samples across 3 independent cohorts. Our analysis leveraged the biological, clinical, and technical heterogeneity across these 37 cohorts to demonstrate that a conserved host response to viral infection is (1) associated with severity, (2) predominantly driven by myeloid cells, and (3) defines distinct trajectories for mild or severe outcomes in patients with viral infection. Using these trajectories, we showed that increased hematopoiesis, myelopoiesis and MDSCs, and reduced NK and T cells are associated with increased severity across all viral infections. Importantly, trajectory analysis identified four gene modules, two of which are associated with a detrimental response leading to a severe outcome, and the other two with a protective response leading to mild infection. Finally, we defined the SoM score using these modules that accurately distinguished patients with mild or moderate viral infection from those with severe outcomes.

Our analysis provides strong evidence of a conserved host immune response to several viruses that is associated with severity. Although we had identified the MVS by analyzing three respiratory viruses (influenza, RSV, and HRV), it is generalizable across novel viruses, including SARS-CoV-2, chikungunya, and Ebola. Our results also demonstrate that the conserved host response to viral infection is generalizable across ages. Out of 37 cohorts, 12 cohorts consisted of 931 samples from children (<18 years), most of which (643 samples in 6 cohorts) were children younger than 2 years. Arguably, this conserved host immune response to viral infection is expected. However, while the majority of the research on the host immune response to SARS-CoV-2 has focused on understanding its dysregulation, how it differs from other viruses, and its association with severity, our results demonstrate a conserved similarity in the dysregulation of the host immune response in patients with severe outcomes, irrespective of the infecting virus. Our findings present several opportunities to improve global pandemic preparedness for the pandemics that will invariably come in the future, including the development of novel diagnostic and prognostic tests, identification of novel drug targets for host-directed broad-spectrum anti-viral therapies, and drug repurposing.

Unlike the MVS score, the SoM score distinguished patients with a severe outcome from those with a non-severe outcome with very high accuracy. This clinically meaningful increase in accuracy of the SoM score is due to the four gene modules associated with either a detrimental or protective host response to viral infection. In contrast, the MVS score considers all genes equal irrespective of their protective or detrimental role. Arguably, the 42-gene signature is not optimal for clinical translation. However, such a conserved gene signature, identified using a large amount of heterogeneous data across multiple cohorts, can be further analyzed to identify a parsimonious, clinically useful, point-of-care test that is generalizable across patient populations. For example, we have previously described a 3-gene host response-based signature for diagnosis of tuberculosis that has been shown to be generalizable across a large number of patient populations, and has been translated into a proof-of-concept point-of-care cartridge with high accuracy (Sodersten et al., 2020). In addition, given the high pairwise correlation between genes within each module, only a small subset of genes within each module would provide the same discriminatory power, further allowing the selection of a parsimonious gene signature. Importantly, trajectory analysis suggests that the SoM score has the potential to predict the severity of outcome in patients with viral infection, though it needs to be tested in additional cohorts. During viral outbreaks or busy winter seasons, when a substantial number of patients are clinically borderline, a test that could improve our ability to accurately distinguish those who can safely recover at home from those who are at high risk of deteriorating would be an indispensable tool to avoid overwhelming the healthcare system. Importantly, this test could also be used for identifying patients at high risk of severe outcomes in clinical practice outside of outbreaks.

Module 3 included a monocyte chemoattractant (*CCL2*), a regulator of type I interferon transcription (*MAFB*), interferon-induced genes (ISGs; *OASL, UBE2L6*), and genes involved in cell death (*TMEM123, CASP7*). These genes were over-expressed in patients with mild viral infection compared to healthy controls and those with severe viral infection, and highly correlated with each other in patients with mild viral infection, but not in those with severe viral infection, irrespective of virus, suggesting that a highly coordinated immune response between monocyte recruitment, interferon response and cell death is associated with protection. Our results are consistent with recent observations that ISGs are strongly induced in patients with moderate SARS-CoV-2 infection compared to those with severe SARS-CoV-2 infection (Arunachalam et al., 2020; Hadjadj et al., 2020), and generalize to patients with non-severe viral infection compared to those with severe infection, irrespective of the virus.

Importantly, the genes in module 3 were more correlated with multiple interferon-induced transmembrane proteins (IFITMs) in patients with mild infection compared to those with severe viral infection. IFITMs are involved in restricting viruses at various stages of the life cycle, including (1) blocking host cell entry by trapping virions in endosomal vesicles, (2) inhibiting viral gene expression and protein synthesis, and (3) disrupting viral assembly (Liao et al., 2019; Zhao et al., 2018). The lower correlations between the expression of the IFITM genes and the genes in module 3 strongly suggest that the interferon-induced response is “decoupled” from the protective response in patients with severe viral infection. Understanding the mechanisms underlying this decoupling could lead to targets for host-directed therapy for viral infection.

Analysis of scRNA-seq in 3 independent cohorts and *in silico* cellular deconvolution across 32 cohorts found increased HSPCs in patients with severe viral infection, irrespective of the virus. In contrast, we have previously shown reduced proportions of HSPCs in mild viral infection (Bongen et al., 2018), which may reflect the production of myeloid cells at the expense of the lymphoid compartment to replenish myeloid cells during infection (Takizawa et al., 2012). Indeed, we observed increased myeloid cells and reduced lymphoid cells in both scRNA-seq and *in silico* cellular deconvolution analysis. This result further supports a model where human HSPCs take an active role in the immune response by differentiating into myeloid cells, similar to what we have previously observed (Bongen et al., 2018). Increased HSPC proportions in patients with severe viral infection suggests that emergency hematopoiesis is associated with increased risk of severity, irrespective of the virus, possibly as the host immune response fails to adequately respond to the infecting virus.

The MVS is predominantly expressed in the myeloid cells. First, we found the MVS increased at a single-cell level in CD14+ monocytes, which increase with the severity of viral infection, whereas CD16+ monocytes decrease, which is in line with several recent studies of patients infected with SARS-CoV-2 (Gatti et al., 2020; Hadjadj et al., 2020; Silvin et al., 2020; Zhou et al., 2020). This suggests that reduced CD16+ monocytes in peripheral blood, possibly due to efflux to the site of infection in response to ongoing tissue damage or dysregulated cytokine sensing, is a conserved feature of the host response in severe viral infection across viruses, and may have prognostic significance. In addition, we found increased proportions of PMN- and monocytic-MDSCs, and anti-inflammatory macrophages along with higher expression of their phenotypic and functional markers in patients with severe viral infection, irrespective of the virus. Interestingly, in patients with mild viral infection, markers of MDSCs did not increase substantially, and the proportions of anti-inflammatory macrophages decreased. These results suggest that lower myeloid-derived suppression in the early phase of infection is protective. These results provide strong evidence that, although increased PMN- and M-MDSCs may limit hyperinflammation during active viral infection, they may lead to a detrimental amplification of immunosuppression, irrespective of the virus. The modulation of monocyte responses, as reflected by gene expression, is compatible with recent findings showing the detrimental role of monocytes/macrophages in severe SARS-CoV-2 infection associated with respiratory dysfunction. Severe respiratory failure in these patients is either driven by macrophage activation syndrome or by a unique pattern of monocyte dysregulation where the expression of HLA-DR is down-regulated while the potential for high cytokine production is maintained (Giamarellos-Bourboulis et al., 2020).

Among their immunosuppressive roles, MDSCs are known to suppress NK cell activity through arginase and ROS/RNS (Schrijver et al., 2019). Indeed, our trajectory and *in silico* deconvolution analyses and scRNA-seq data found several NK cell-specific genes (*KLRB1, KLRG1, KLRD1, PIK3R1)* were negatively correlated with the severity of viral infection, and the proportions of NK cells reduced in patients with severe viral infection. We have previously shown that healthy individuals with lower expression of *KLRD1* are more likely to be infected when challenged (Bongen et al., 2018). A negative correlation between expression of *KLRD1* and the severity of viral infection, including SARS-CoV-2, further emphasizes that *KLRD1*-expressing NK cells may play a protective role following infection, irrespective of the infecting virus.

Taken together, our analyses offer a systems view of the immune state during viral infection and factors that mediate and predict progression to mild or severe outcomes, irrespective of the clinical, biological, and technical heterogeneity and the infecting virus. Our findings identified host response modules that could lead to new intervention strategies, including diagnostics for predicting patients at higher risk of severe outcomes, and broad-spectrum host-directed therapies for improved pandemic preparedness.

## Methods

### Dataset collection and preprocessing

We downloaded 26 gene expression datasets from the National Center for Biotechnology Information (NCBI) Gene Expression Omnibus (GEO), Sequence Read Archive (SRA), ArrayExpress, and European Nucleotide Archive (ENA), consisting of 4,780 samples across 34 independent cohorts derived from whole blood or peripheral blood mononuclear cells (PBMCs) (**Table S1**). The samples in these datasets represented the biological and clinical heterogeneity observed in the real-world patient population, including healthy controls and patients infected with 16 different viruses with severity ranging from asymptomatic to fatal viral infection over a broad age range (<12 months to 73 years) (**Figure 1A** and **Table S1**). Notably, the samples were from patients enrolled across 18 different countries representing diverse genetic backgrounds of patients and viruses. Finally, we included technical heterogeneity in our analysis as these datasets were profiled using microarray and RNA sequencing (RNA-seq) from different manufacturers.

We renormalized all microarray datasets using standard methods when raw data were available from the GEO database. We applied GC robust multiarray average (gcRMA) to arrays with mismatch probes for Affymetrix arrays. We used normal-exponential background correction followed by quantile normalization for Illumina, Agilent, GE, and other commercial arrays. We did not renormalize custom arrays and used preprocessed data as made publicly available by the study authors. We mapped microarray probes in each dataset to Entrez Gene identifiers (IDs) to facilitate integrated analysis. If a probe matched more than one gene, we expanded the expression data for that probe to add one record for each gene. When multiple probes mapped to the same gene within a dataset, we applied a fixed-effect model. Within a dataset, cohorts assayed with different microarray types were treated as independent.

### Standardized severity assignment

For each dataset, we used the sample phenotypes as defined in the original publication. We manually assigned a severity category to each sample based on the cohort description for each dataset in the original publication as follows: (1) healthy controls – asymptomatic, uninfected healthy individuals, (2) asymptomatic or convalescents – afebrile asymptomatic individuals who tested positive for a virus or those fully recovered from a viral infection with completely resolved symptoms, (3) mild – symptomatic individuals with viral infection that were either managed as outpatient or discharged from the emergency department (ED), (4) moderate – symptomatic individuals with viral infection who were admitted to the general wards and did not require supplemental oxygen, (5) serious - symptomatic individuals with viral infection who were described as ‘severe’ by original authors, admitted to general wards with supplemental oxygen, or admitted to the intensive care unit (ICU) without requiring mechanical ventilation or inotropic support, (6) critical - symptomatic individuals with viral infection who were on mechanical ventilation in the ICU or were diagnosed with acute respiratory distress syndrome (ARDS), septic shock, or multiorgan dysfunction syndrome (MODS), and (7) fatal – patients with viral infection who died in the ICU.

For datasets that did not provide sample-level severity data (GSE101702, GSE38900, GSE103842, GSE66099, GSE77087), we assigned severity categories as follows. We categorized all samples in a dataset as “moderate” when either (1) >70% of patients were admitted to the general wards as opposed to discharged from the ED, (2) <20% of patients admitted to the general wards required supplemental oxygen, or (3) patients were admitted to the general wards and categorized as ‘mild’ or ‘moderate’ by the original authors. We categorized all samples in a dataset as “severe” when >20% of patients had either (1) been admitted to the general wards and categorized as ‘severe’ by original authors, (2) required supplemental oxygen, or (3) required ICU admission without mechanical ventilation.

### Viral challenge studies

GSE73072 included seven viral challenge studies that determined the infection status of a subject through reverse transcription PCR (RT-PCR) for a given virus (H1N1, H3N2, RSV, HRV) in longitudinally collected nasopharyngeal samples. In these studies, we assigned all baseline pre-challenge samples and subjects who never shed virus, as determined by RT-PCR, to the ‘healthy’ category. We assigned samples from infected subjects, defined as those who had virus detected in any of their nasopharyngeal samples, to one of three categories: (1) before infection - blood samples collected after challenge but before a virus was detected in a nasopharyngeal sample, (2) after infection - blood samples collected after the last nasopharyngeal sample in which a virus was detected, and (3) during infection - blood samples collected between the first and last nasopharyngeal sample in which a virus was detected.

### COCONUT co-normalization

We used Combat CONormalization Using conTrols (COCONUT) for between-dataset normalization (Sweeney et al., 2016b). COCONUT allows for co-normalization of gene expression data without bias towards sample diagnosis by applying a modified version of the ComBat empirical Bayes normalization method (Johnson et al., 2006), which assumes a similar distribution between control samples. Briefly, healthy controls from each cohort undergo ComBat co-normalization without covariates, and the ComBat estimated parameters are computed for healthy samples in each dataset. By applying these parameters to the non-healthy samples, all datasets keep the same background distribution while retaining the same relative distance between healthy and disease samples, which preserves the biological variability between the two groups within a dataset. We have previously shown that when COCONUT co-normalization is applied, housekeeping genes remain invariant across both conditions and cohorts, and each gene retains the same distribution across conditions within each dataset (Sweeney et al., 2016b).

### MVS genes and score

We did not derive a *de novo* gene signature to represent a conserved host response to viral infection. Instead, we used our previously described 396-gene signature from peripheral blood (Andres-Terre et al., 2015). Further, as previously described, we defined the MVS score of a sample as the difference between the geometric mean of the over-expressed genes and the geometric mean of the under-expressed genes in the MVS (Andres-Terre et al., 2015). Out of 396 genes in the MVS, 251 genes (111 over- and 140 under-expressed) were measured across all datasets. Across 4 independent datasets that measured all 396 genes in patients infected with SARS-CoV-2, Ebola, or chikungunya, we found that the MVS score using the 251-gene signature was highly correlated with the MVS score using the 396-gene signature (0.976≤r≤0.997; **Figure S7**). Thus, the 251-gene signature provided the same information and did not skew our results (**Figure S7**). We measured the correlation of the MVS score with viral infection severity using Spearman’s rank correlation coefficient. We used the Mann–Whitney U test (Wilcoxon rank-sum test) to compare MVS scores between two groups. We tested the trend of the MVS score along viral infection severity categories using the Jonckheere-Terpstra trend test.

### RNA sequencing analysis

We obtained the raw reads for the Ebola (PRJNA352396) and chikungunya (PRJNA507472 and PRJNA390289) cohorts from the European Nucleotide Archive (ENA). We obtained the RNA-seq raw reads of the SARS-CoV-2 cohort from Inflammatix. We assessed the quality of the raw reads with Trim Galore (v0.6.5), trimmed Illumina adaptors, and removed reads that were too short after adaptor trimming (less than 20 nt). We then mapped the cleaned reads to human genome sequences (hg38) using STAR (v2.7.3) (Dobin et al., 2013). We performed more quality control by checking the quality of the mapped reads in BAM format with Qualimap (v.2.2.2) (García-Alcalde et al., 2012). To quantify gene expression, we obtained human transcriptome sequences from GENCODE site (v32), then processed the cleaned reads with Salmon (1.2.1) (Patro et al., 2017) to get transcript-level expression. Using Tximport (v1.16.0) (Soneson and Robinson, 2018), we then summarized to gene-level expression. Finally, we applied the variance stabilizing transformation from DESeq2 (v1.26.0) (Love et al., 2014) to normalize gene expression for downstream analysis and visualization.

### Detection of viral reads in RNA-seq data

We obtained genome sequences of 501 human viruses from the NCBI virus database (accessed on April 19, 2020). We concatenated the list of viral sequences with the list of human transcriptome sequences and then built a decoy-aware index using Salmon. We mapped the reads to the concatenated index using Salmon with a selective-alignment algorithm, which, together with the decoy-aware index, mitigates potential spurious mapping of reads arising from unannotated human genomic loci and reduces false positives. We extracted reads mapped to viral genomes and filtered them to remove secondary alignments and paired-end reads with only one mate mapped. We also checked the reads with NCBI Nucleotide BLAST to ensure viral origin. We normalized the viral read counts by the total number of sequencing reads of each sample. We measured the correlation between the MVS score and viral read counts using Pearson correlation coefficient.

### Analysis of single-cell RNA-seq data

We downloaded the scRNAseq data for (1) the Stanford Cohort from the COVID-19 Cell Atlas (Wilk et al., 2020), and (2) the Atlanta cohort from the NCBI GEO (Arunachalam et al., 2020). We processed scRNAseq data for the Seattle cohort (Su et al., 2020) with Cell Ranger (v3.1.0) (Zheng et al., 2017) and performed quality control on the three datasets with Seurat (Satija et al., 2015). We normalized read counts using regularized negative binomial regression with the ‘SCTransform’ function. Then we applied integration workflow in Seurat to integrate the three datasets using canonical correlation analysis. We performed Principal component analysis (PCA), Uniform Manifold Approximation and Projection (UMAP), and Shared Nearest Neighbors clustering on the integrated expression data. Cell type annotation of clusters was performed with both SingleR (Aran et al., 2019) and manual annotation using cell type markers.

### *In silico* cellular deconvolution using immunoStates and multi-cohort analysis of estimated cellular proportions

We performed *in silico* cellular deconvolution using immunoStates as a basis matrix with support vector regression to estimate proportions of 25 immune cell subsets in each sample (Vallania et al., 2018).

To investigate changes in the immune cell proportions between patients with different severity of viral infection, we conducted three multi-cohort analyses using MetaIntegrator R package (Haynes et al., 2017) between samples from the following categories: 1) subjects with non-severe viral infection (severity categories ‘mild’ and ‘moderate’) vs healthy controls, 2) subjects with severe viral infection (severity categories ‘serious’, ‘critical’, and ‘fatal’) vs healthy controls, and 3) subjects with severe viral infection vs subjects with non-severe viral infection (**Table S3**). We combined effect sizes across studies using a random-effects inverse variance model. For each meta-analysis, we calculated the change in proportions for each immune cell type between groups in each cohort as the Hedges’ g effect size (ES). We corrected p-values for multiple hypotheses testing using the Benjamini-Hochberg correction to obtain the false discovery rate (FDR). We used a threshold of FDR < 20% and representation in a minimum of 5 studies in conjunction with leave-one-out analysis to identify immune cell types with increased or decreased proportions between groups. Individual samples that met the following criteria were excluded: non-viral infection, non-healthy controls, and one sample from PRJNA252396 (SRR4888654) which had the same expression value for all 317 genes. Datasets with less than two samples in each of the compared groups were excluded from meta-analysis.

### Trajectory inference analysis

We co-normalized 1674 samples from 21 cohorts in 19 datasets with 1509 samples from four independent challenge studies using COCONUT. Each challenge study inoculated healthy volunteers with one of four viruses (HRV, RSV, H1N1, and H3N2). We adapted tSpace, a method for identifying cellular differentiation trajectories using scRNA-seq data (Dermadi et al., 2020), to identify disease trajectories using bulk transcriptome microarray profiles. We refer to the adaption to bulk transcriptome data as disease space (dSpace), although the core method remains identical to tSpace. The tSpace algorithm involves three steps: (1) calculation of a set of sub-graphs, (2) calculation of the trajectory space matrix across the sub-graphs, and (3) visualization. In the first step, we calculated a set of sub-graphs keeping L out of K nearest neighbors in a KNN graph. The user defines the number of sub-graphs (G), neighborhood size (K), and how many nearest neighbors will be preserved in the sub-graphs (L). The second step computes a *trajectory space* distance matrix using a modified Dijkstra algorithm that implements waypoints (WP) to exponentially weigh and refine distances. The final *trajectory space* matrix is a dense matrix in which each sample is a row, and calculated trajectories are columns. The number of trajectories (T > 150) is user-defined and very robust across a wide dynamic range. Finally, we visualize the samples and their relationships in trajectory space using PCA or UMAP.

We used the following parameters for the dSpace analysis: G=5, K=65, L = 49, T=500, WP=20. We used Pearson correlation as the metric for computing distance between two samples. We fitted a principal line through data visualized in the first two components of tSpace (tPC1, tPC2) using the princurve R package. Princurve calculates lambda, an arc length distance for each data point, which we used to align subjects along the isolated trajectory. Furthermore, the covariance matrix of the transposed trajectory matrix (covariance mapping) coupled with the hierarchical clustering identified clusters of patients with shared trajectory space. The covariance matrix of the transposed trajectory matrix allows identification of patients that belong to diverging trajectories, and hierarchical clustering of covariance matrix allowed us to group patients that are in severe and non-severe branches, thus enabling isolation of both branches. Each of the determined clusters is a reflection of position of patients in the trajectory space. Hierarchical clustering was calculated using hclust and Dist R functions with “euclidean” and “complete” parameters.

Severe and non-severe branches shared a substantial number of healthy patients. Therefore, we aligned them using dynamic time warping (dtw R package) and split them into 4 stages. All 251 genes and the fitted trajectory (lambda value) were used for alignment. We applied a permutation test (Efron and Tibshirani, 2002) for each of the 4 stages and identified 96 genes that were differentially expressed within the same stage between the two severity branches. In our testing we used 1000 permutations, and for significance FDR < 0.001 and |effects size| > 0.3.

### Calculation of the SoM score

The Severe or Mild (SoM) score is a 42-gene model that utilizes the expression of genes from the 4 gene modules to distinguish between severe and mild viral infection. For each sample, we compute the geometric mean of the expression of genes from each module. Then, we calculate a score by taking the sum of the geometric means of modules 1 and 2 and dividing that by the sum of the geometric means of modules 3 and 4, as shown in the following equation:

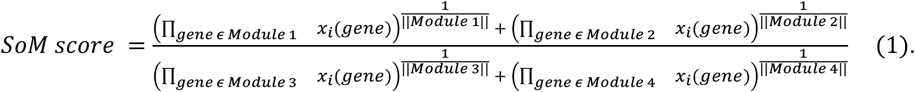

## Supporting information

Summary description of datasets used in the analyses

## Data Availability

All data are publicly available from public repositories of gene expression data including the Gene Expression Omnibus, EBI ArrayExpress, Sequence Read Archive, and others. The identifiers for each study are listed in Supplementary Table 1 and all figures.

## Author contributions

PK conceived and supervised the study. HZ, AMR, DDB, JT, LMJ, and MD collected, annotated, processed, and analyzed data. PK, HZ, AMR, DDB, JT, LMJ, MD, and YL interpreted analyses results. PK, HZ, AMR, DDB, JT, LMJ, MD, and YL wrote the manuscript. MK, TM, and EJGB enrolled patients with SARS-CoV-2 infection in Greece. YHB and YDH profiled and processed RNA sequencing data from the patients with SARS-CoV-2 infection in Greece. YS and JH profiled single-cell RNA-seq of PBMCs from patients with SARS-CoV-2 infection in the Seattle Cohort.

### Acknowledgments

PK is funded in part by the Bill and Melinda Gates Foundation (OPP1113682); the National Institute of Allergy and Infectious Diseases (NIAID) grants 1U19AI109662, U19AI057229, and 5R01AI125197; Department of Defense contracts W81XWH-18-1-0253 and W81XWH1910235; and the Ralph & Marian Falk Medical Research Trust. AMR and YL are funded by National Science Foundation Graduate Research Fellowship. AMR is also funded by Stanford Graduate Fellowship. YL is also funded by the Knight-Hennessy Scholars Program. JT is funded by National Science Scholarship (PhD) from the Agency of Science, Technology, and Research (A*STAR), Singapore.

## Supplementary figure legends

**Figure S1: MVS score is associated with severity of viral infection in each dataset in discovery and validation cohorts. a)** ROC curves for the MVS score in each dataset in the discovery cohorts. AUROC values varied from 0.859 (95% CI 0.69-1) to 1 (95% CI 1-1). **b)** ROC curves for the MVS score in 4 independent cohorts profiled using RNAseq. AUROC values varied from 0.84 (95% CI 0.76-0.92) to 0.972 (95% CI 0.932-1). **c**) Violin plots of the MVS score for all samples across 19 datasets.

**Figure S2: MVS score is predominantly from the myeloid cells and Neutrophil proportion is higher in patients with severe viral infection. a)** MVS score for cell types in each sample of the three scRNA-seq cohorts. Red color indicates high score and blue indicates low score. The size of the circle is proportional to the variability of the MVS score across all cells of a specific cell type in a sample. The bar plot on the right of this panel shows the cell type proportions of each sample. **b)** Forest plots for the comparisons between proportions of neutrophils in bulk transcriptomic profiles. Each row represents an effect size of the comparison of non-severe patients vs healthy controls (top), severe patients vs healthy controls (center), and severe vs non-severe patients (bottom).

**Figure S3: Samples from viral challenge studies almost exclusively cluster with samples from non-severe viral infection within dSpace. a)** dSpace trajectory analysis of 3,183 samples from 25 cohorts, including 1,509 samples from 4 viral challenge studies (2 influenza, 1 HRV, 1 RSV). Each point represents a sample; viral challenge study samples are demarcated as triangles. **b)** Proportion of samples for each severity category or challenge study group within each cluster.

**Figure S4: Expression levels of genes associated with NK cells, myeloid cell-derived suppression, HSPCs, and an overall protective host response correlate with severity of viral infection. a-b)** *KLRD1* and *PIK3R1* expression levels in **a)** the discovery cohort and **b)** SARS-CoV-2 infection. **c-d)** Expression levels of myeloid cell-associated genes, including MDSC markers and *ORM1* in **c)** the discovery cohort and **d)** SARS-CoV-2 infection. **e-f)** Expression levels of genes over-expressed in patients with severe viral infection but not in those with non-severe viral infections compared to healthy controls, and preferentially expressed in circulating HSPCs in **e)** the discovery cohort and **f)** SARS-CoV-2 infection. **g-h)** Expression levels of genes identified to be significantly higher in patients with mild viral infection compared to those with serious, critical, or fatal viral infection, or healthy controls, in **g)** the discovery cohort and **h)** SARS-CoV-2 infection.

**Figure S5: The interferon-induced genes (*IFITM1, IFITM2, IFITM3)* and type I and II interferon receptors are highly correlated with protective response genes in mild but not severe viral infection. a)** Expression of *IFITM1, IFITM2, IFITM3*, and type I and II interferon receptors across severity categories in patients with different viral infections including SARS-CoV-2. Each point in the violin plots represents a sample. **b)** Boxplots representing the correlation between IFITMs and type I and II interferon receptors, and the genes belonging to the protective response module. Each point represents a correlation between a gene pair, and lines connect the same pair across severity categories. P-values for the comparison between severity categories were computed using Wilcoxon signed-rank test.

**Figure S6: SoM score distinguishes mild and severe viral infection in discovery and validation cohorts with higher accuracy than the MVS score. a-b**) The ROC curves of the MVS score for differentiating between severity categories of viral infection in **a)** discovery and **b)** validation cohorts. **c-d**) The ROC curves of the SoM score for differentiating between severity categories of viral infection in **c)** discovery and **d)** validation cohorts.

**Figure S7: The reduced 251-gene MVS score is highly correlated with the 396-gene MVS score**. Scatter plots comparing the MVS score of each sample in 4 RNA-seq cohorts computed using the full set of 396 genes (X axis) with the score computed using the reduced set of 251 genes that were measured in the majority of the discovery datasets.

