## Supplementary figures and images for "Multi-cohort analysis of host immune response identifies conserved protective and detrimental modules associated with severity irrespective of virus"

### Supplementary figures

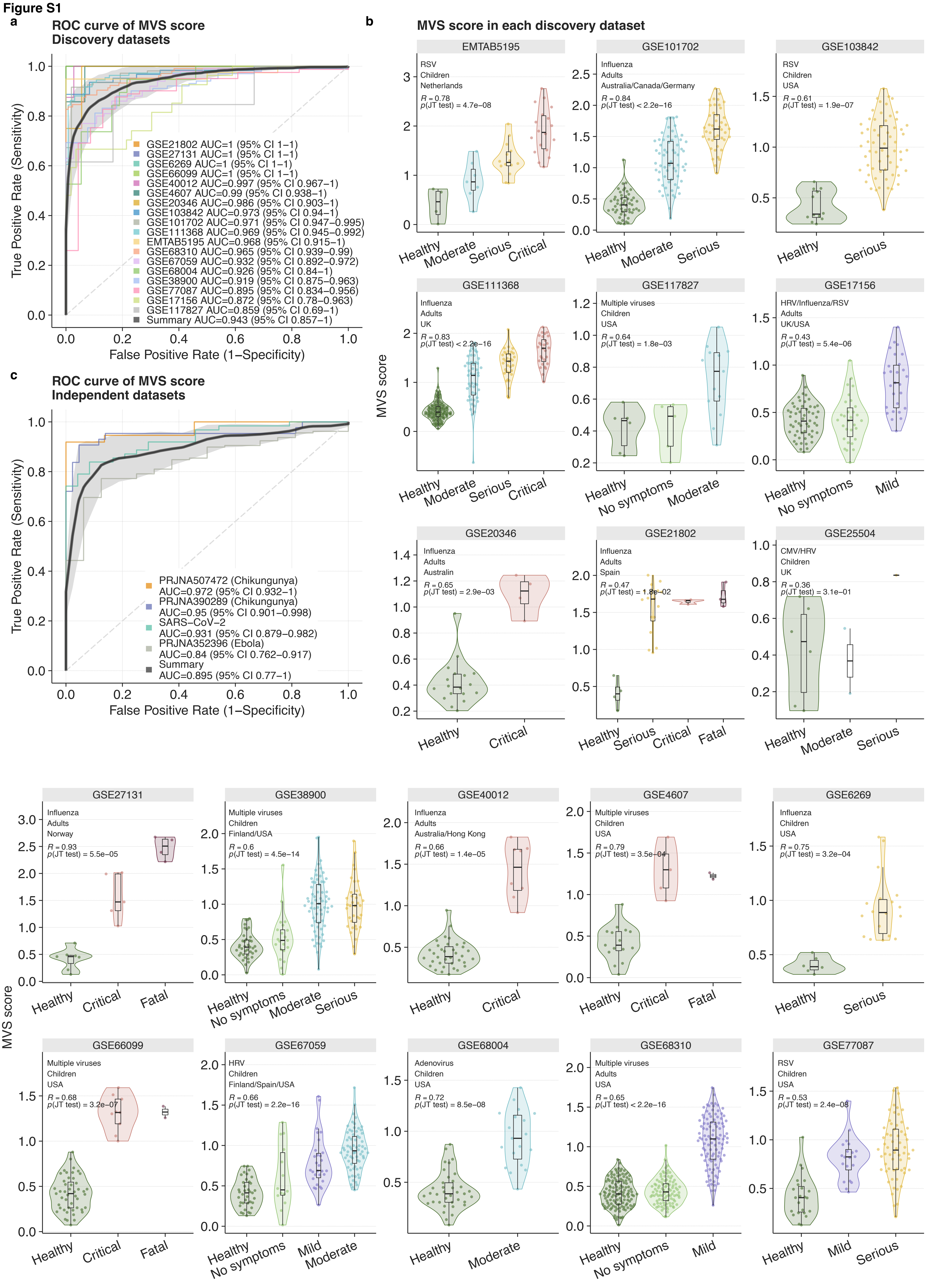

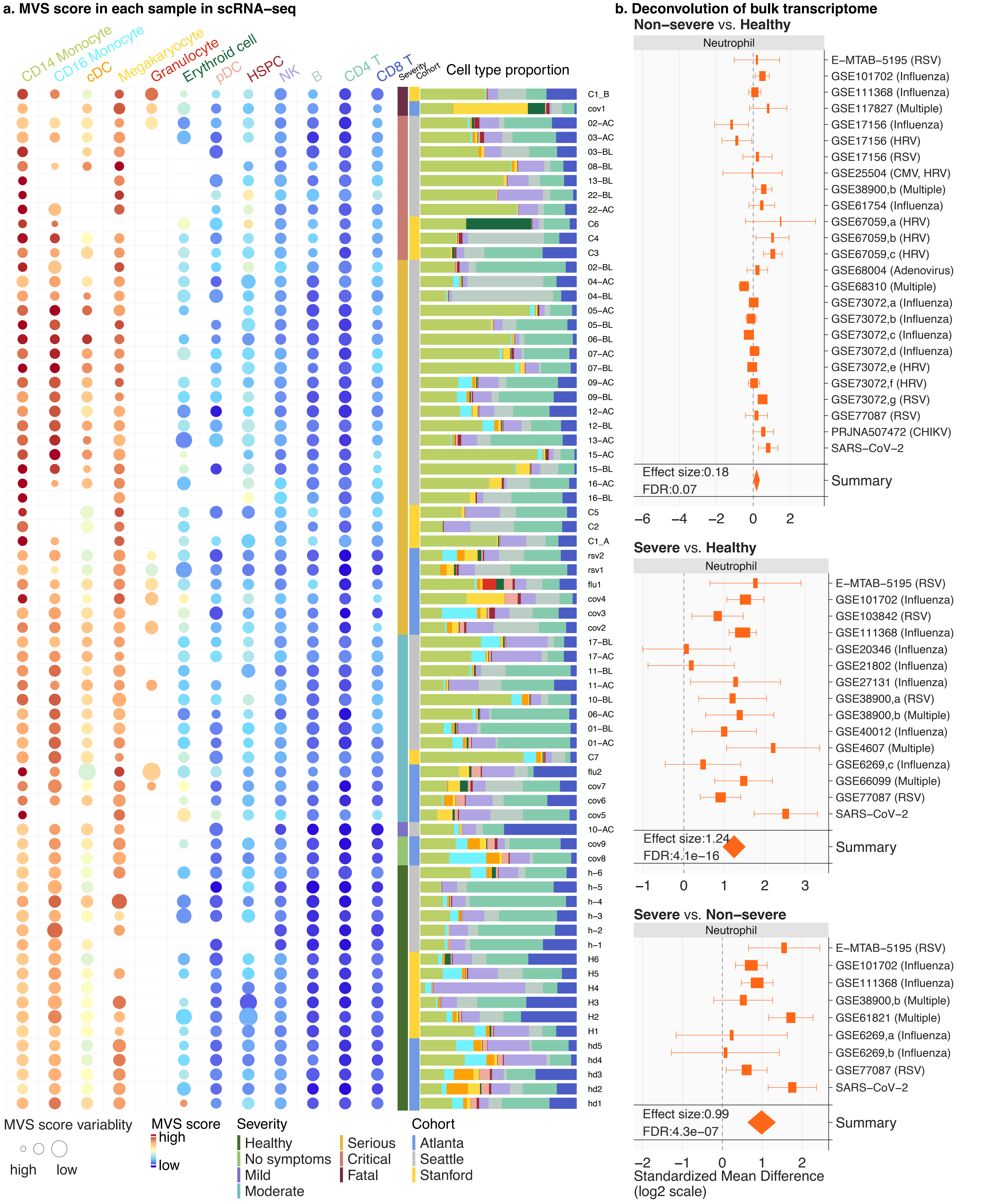

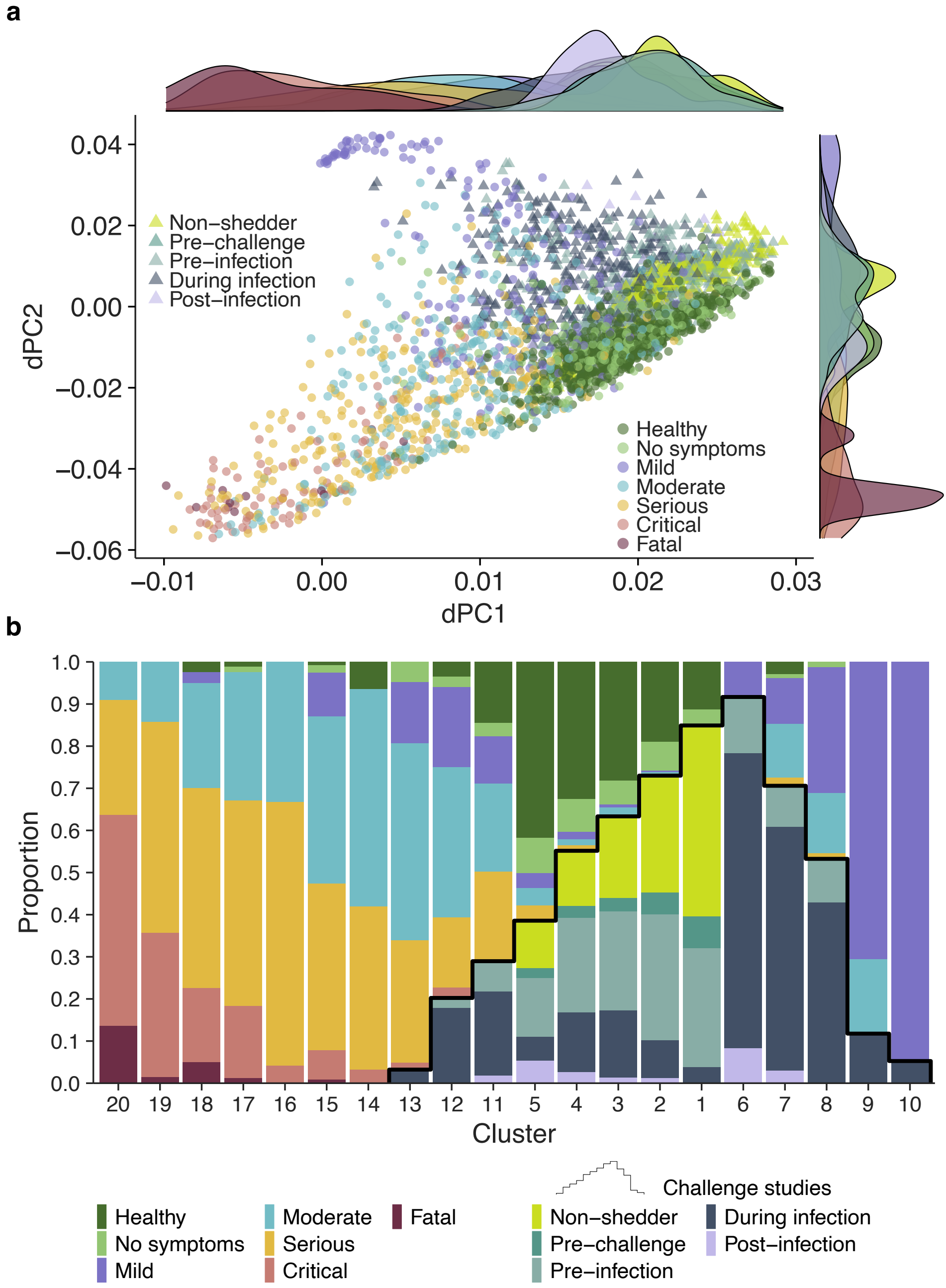

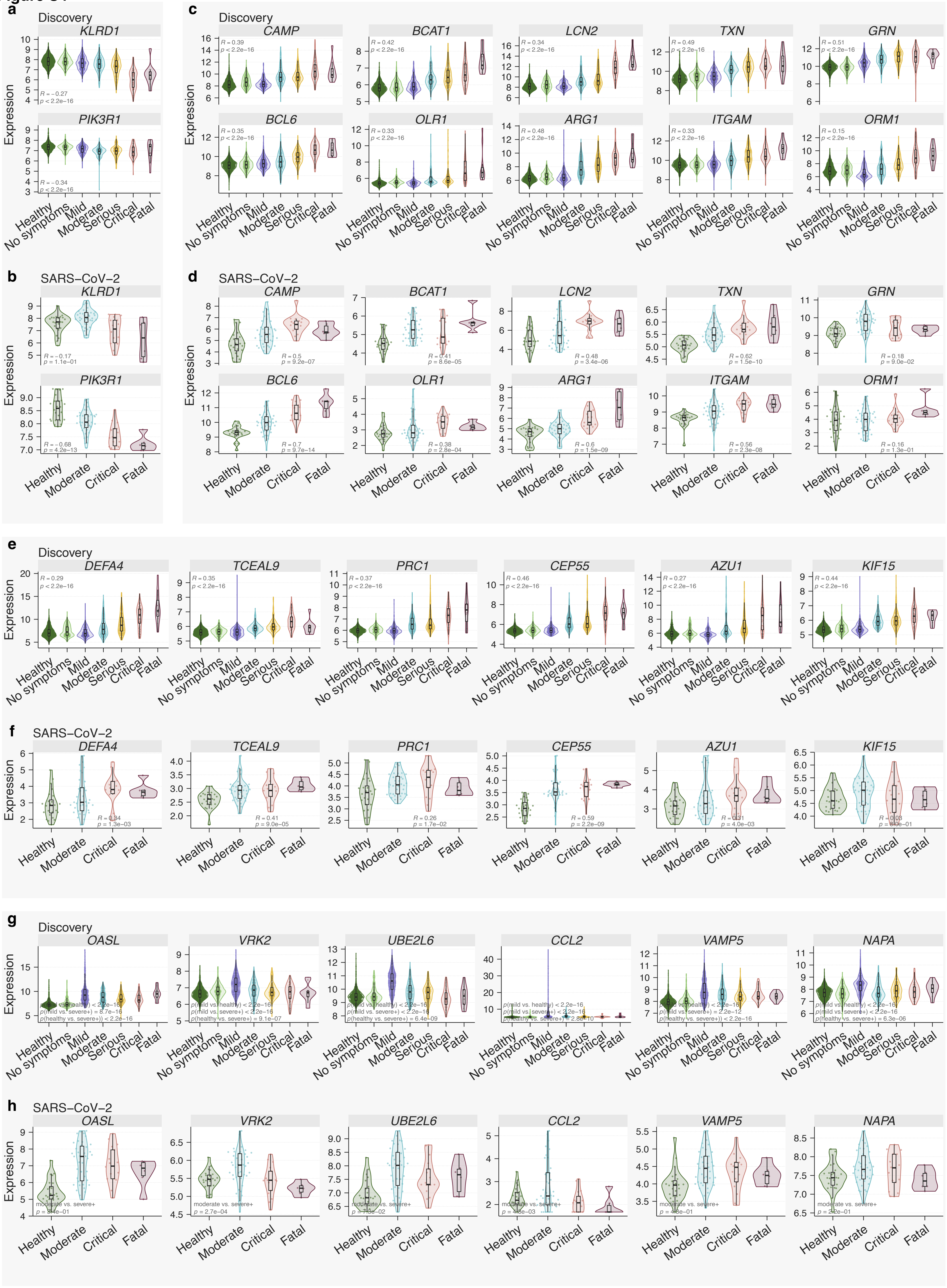

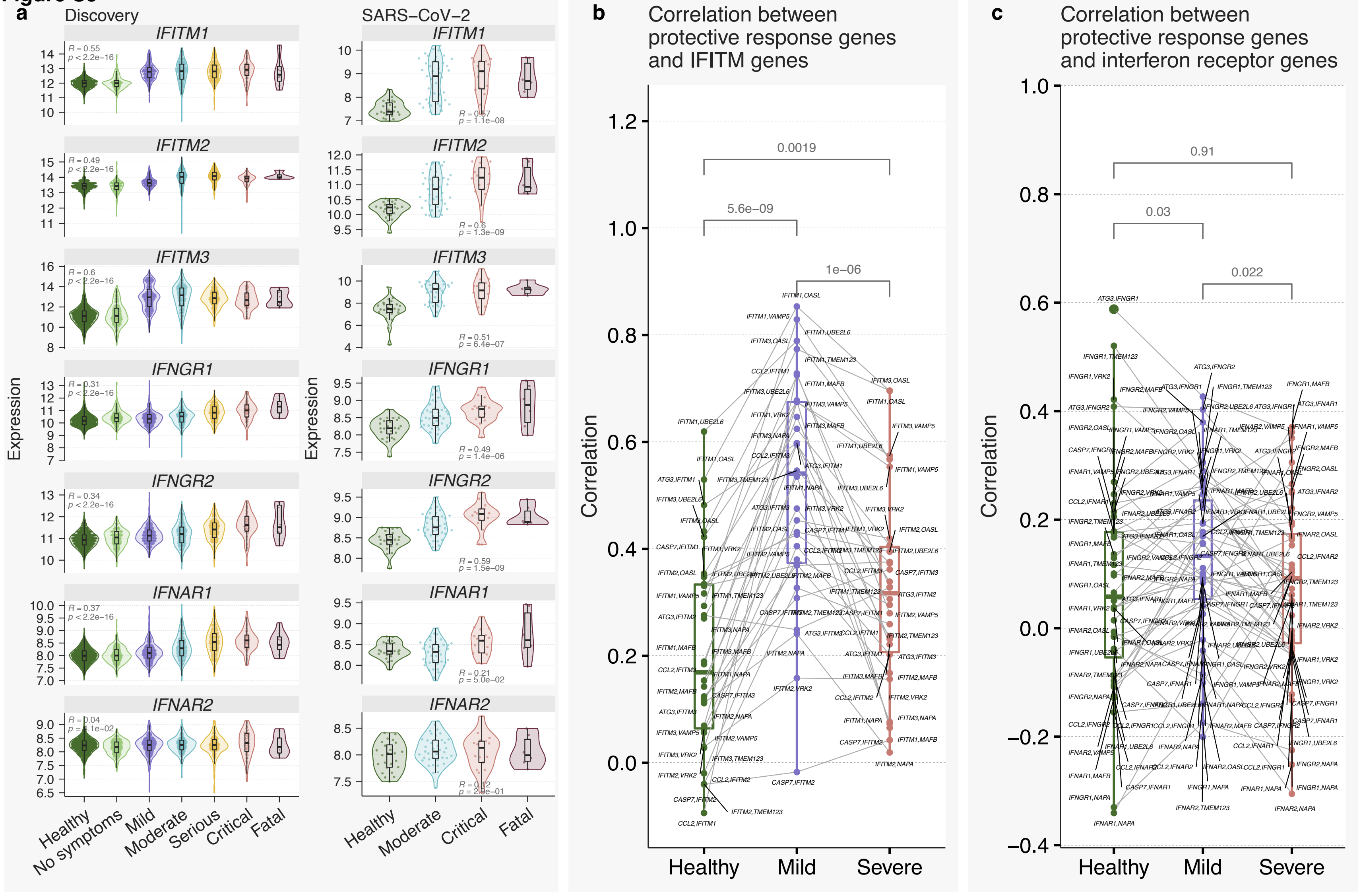

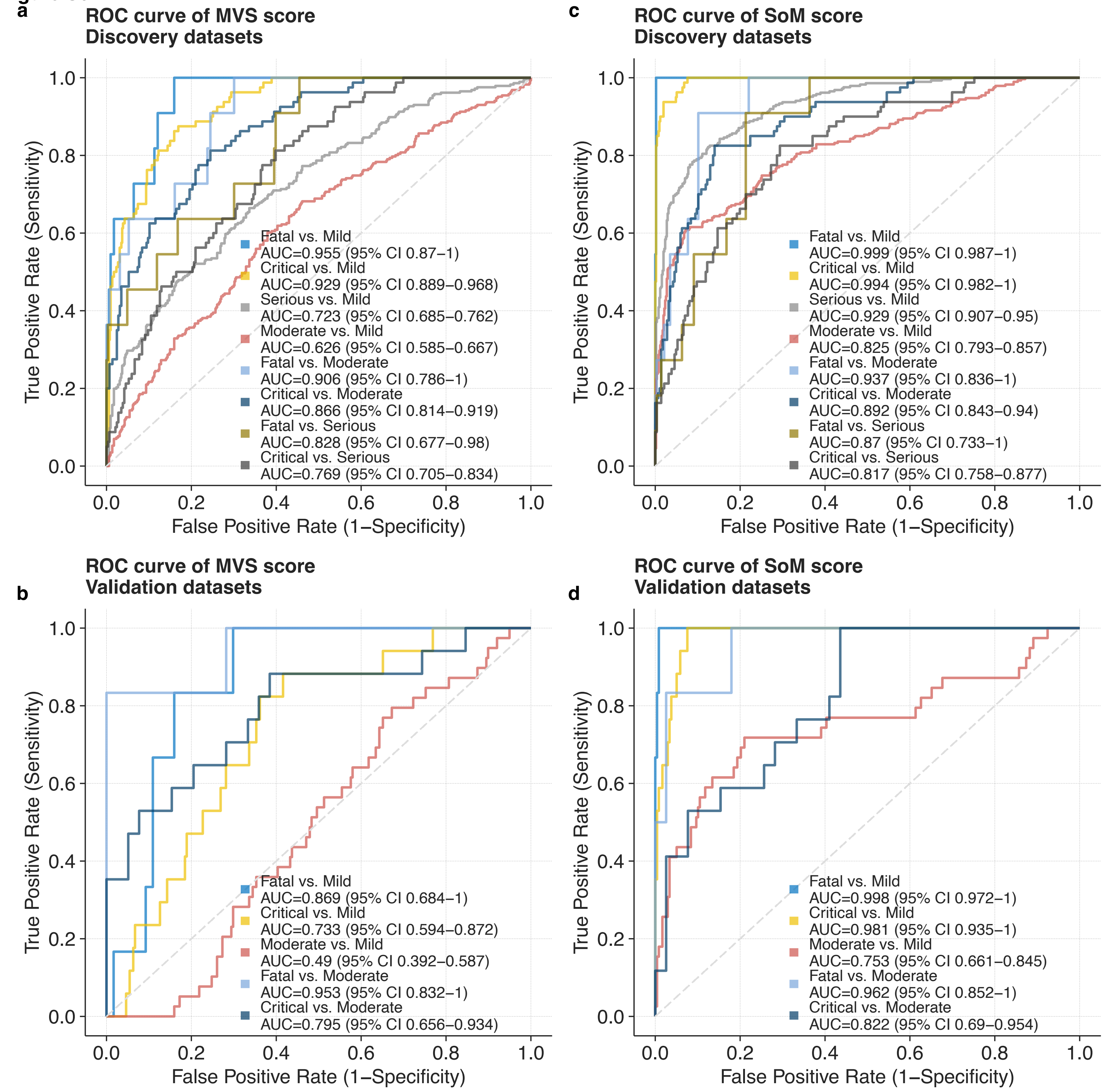

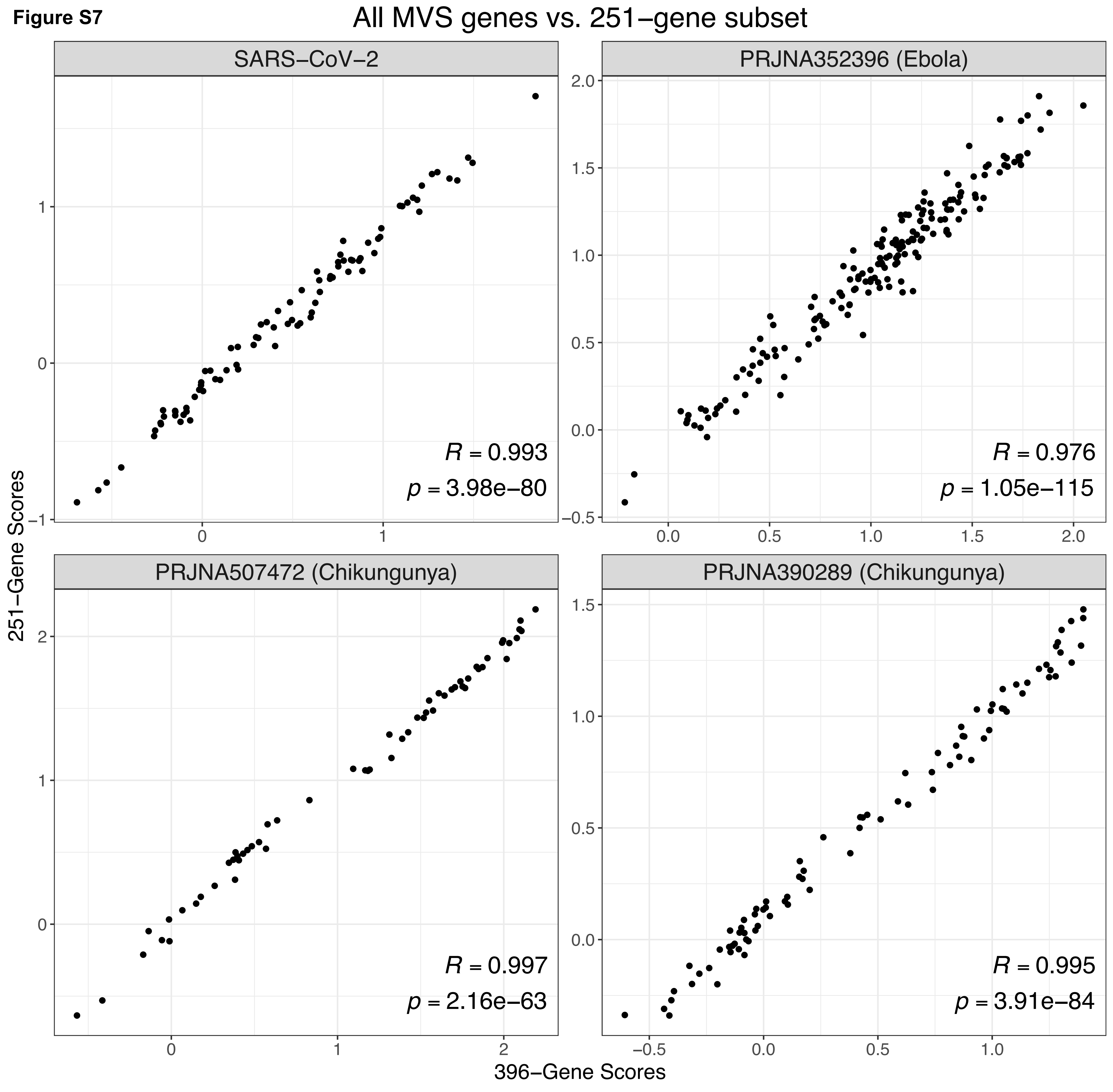
