## Supplementary material for "Multi-cohort analysis of host immune response identifies conserved protective and detrimental modules associated with severity irrespective of virus": Summary description of datasets used in the analyses

**Study Summaries**

This section describes each of the datasets used in the analyses. In order to provide the most accurate information, their descriptions have been used verbatim from the original corresponding manuscripts as much as possible.

**PRJNA352396:** Liu *et al.* (Liu et al., 2017) collected peripheral blood from infected and convalescent, recovering Ebola virus (EBOV) patients located in Guinea during the 2013-2016 outbreak. They first used deep sequencing to define the transcriptomic profile of blood taken from acute patients who either went on to survive or die from EBOV. These transcriptional signatures from acutely ill patients were compared to profiles obtained from former patients who had recovered from EBOV and were EBOV-negative by qRT-PCR and also to data obtained from healthy volunteers mined from historical datasets.

**PRJNA507472:** Soares *et al.* (Soares-Schanoski et al., 2019) collected blood samples from adult subjects reporting arbovirus-like symptoms and controls in Sergipe and São Paulo Brazil. A subset of the patients returned for clinical follow-up, and these Chikungunya (CHIKV) infected patients were under treatment and their joint manifestations were monitored quarterly. Patients who presented joint pain associated or not with joint or periarticular edema at the onset or significant worsening (in cases of previous joint disease) of the acute febrile syndrome after three months were considered chronic. Real-time RT-PCR was performed to test for CHIKV, Zika virus, and Dengue (DENV).

**PRJNA390289:** Michlmayr *et al.* (Michlmayr et al., 2018) selected children (aged 6 months to 14 years) with Chikungunya (CHIKV) infection who were enrolled in the ongoing study at the National Pediatric Reference Hospital (Hospital Infantil Manuel de Jesús Rivera; HIMJR) in Nicaragua. CHIKV cases were laboratory-confirmed by detection of CHIKV using real-time RT–PCR in some cases followed by virus isolation, in acute-phase samples. In addition, seroconversion by IgM capture ELISA and/or a >4-fold increase in antibody titer by Inhibition ELISA in paired acute and convalescent sera were evaluated. Participants were also screened for dengue virus (DENV) infection, and CHIKV/DENV co-infections were excluded. Children with severe symptoms were excluded. All selected cases had an acute-phase sample collected on days 1–2 of illness and a convalescent sample collected on days 15–17 post-illness for serum, whole blood in PAXgene solution, and PBMCs; these were subject to transcriptomic analysis via RNAseq, CHIKV viral titer assays, multiplex ELISA for cytokines, and mass cytometry for cellular phenotyping. Follow-up samples were collected at 3, 6, and 12 months post-infection to study long-term outcomes of CHIKV infection. Sampling times closely adhered to the targeted acute (standard deviation [SD] = 0.5 days) and convalescent (SD = 0.6 days) timepoints. Clinical information was collected every 12 h, and after systematic monitoring by a clinical supervisor was digitized by double-data entry with quality control checks performed daily and weekly.

**SARS-CoV-2:** Giamarellos-Bourboulis *et al.* (Giamarellos-Bourboulis et al., 2020) collected samples from adult patients with community-acquired pneumonia (CAP) and sepsis and CAP by SARS-CoV-2 of either gender within the first 24 h of hospital admission. CAP was defined as the presence of new infiltrate in chest X-ray in a patient without any contact with any health- care facility the last 90 days; sepsis was defined by the Sepsis-3 criteria. COVID was diagnosed for patients with CAP confirmed by chest X-ray or chest computed tomography and positive molecular testing of respiratory secretions. For patients who required mechanical ventilation (MV), blood sampling was performed within the first 24 h from MV and results were used for this analysis. Exclusion criteria were infection by the human immunodeficiency virus; neutropenia; and any previous intake of immunosuppressive medication (corticosteroids, anti-cytokine biologicals, and biological response modifiers). Severe respiratory failure (SRF) was defined as a severe decrease of the respiratory ratio necessitating intubation and MV. All data of patients with bacterial CAP screened for participation in the randomized clinical trial with the acronym PROVIDE (ClinicalTrials.gov NCT03332225) were used. All patients admitted for CAP by SARS-CoV-2 from March 3 until March 30, 2020 participated in the study. RNAseq was performed on PBMCs.

**GSE77087:** De Steenhuijsen Piters *et al.* (de Steenhuijsen Piters et al., 2016) prospectively enrolled previously healthy children (<2 years) with a first episode of RSV infection during four consecutive RSV seasons at Nationwide Children's Hospital, Columbus, Ohio either at the outpatient clinics or within a median of 24 hours (interquartile range 17–39 h) of admission in the pediatric ward or the pediatric intensive care unit (PICU). Asymptomatic healthy control subjects were enrolled during routine primary care visits or elective surgery not involving the respiratory tract. In addition to the need for hospitalization, RSV disease severity was assessed using a clinical disease severity score and by the need for supplemental oxygen, PICU admission, and length of stay. At enrollment, they obtained a blood sample from patients and controls for white blood cell count and transcriptome analysis, a nasopharyngeal bacterial swab for bacterial quantitative polymerase chain reaction (PCR) and microbiome analysis, and a nasal wash for RSV quantitation. RNA was extracted from whole-blood samples and hybridized onto Illumina HT12-V4 beadchips.

**GSE73072:** *Liu et al.* (Liu et al., 2016) quantified the diagnostic advantage of pairing a person's baseline reference with his or her target sample using gene microarray data collected in a large-scale serially sampled respiratory virus challenge study. The data was collected over 7 challenge studies performed by Duke University under a contract awarded by the DARPA Predicting Health and Disease program. These 7 challenge studies are referred to as RSV DEE1, H3N2 DEE2, H1N1 DEE3, H1N1 DEE4, H3N2 DEE5, HRV UVA, and HRV DUKE. These studies consist of 2886 microarray chips assaying 12,023 genes of 148 human volunteer subjects under 4 different inoculation regimes (HRV, RSV, H1N1, H3N2). During each challenge study, subjects had peripheral blood taken prior to inoculation with virus (baseline), immediately prior to inoculation (pre-challenge) and at set intervals following challenge. RNA was extracted at Expression Analysis (Durham, NC) from whole blood using the PAXgene™ 96 Blood RNA Kit (PreAnalytiX, Valencia, CA) employing the manufacturer's protocol. Hybridization and microarray data collection were performed using the Human Genome U133A 2.0 Array (Affymetrix, Santa Clara, CA).

**GSE68310:** Zhai *et al.* (Zhai et al.) enrolled healthy adults aged 18 to 49 to be followed for acute respiratory illness (ARI) through two consecutive influenza seasons 2009-2010 and 2010-2011. Baseline blood specimens were obtained during enrollment and subjects were given thermometers and instructions to call and report for evaluation within 48 hours of ARI onset. Those persons who had a new ARI were seen within 48 hours of onset (day 0) and 2, 4, and 6 days later for repeat evaluation, blood specimen collections, and medical care (including the antiviral zanamivir if indicated) and 21 days later for collection of convalescent specimens. Nasal wash samples were collected for virus detection by RT-PCR on day 0 and day 2. Surveillance for influenza was terminated after 5.5 months; all subjects were asked to return for specimen collection and to provide a medical and ARI history in spring of next year. Serum specimens obtained at baseline, day 0 and day 21 visits for illnesses, and the terminal visit were tested simultaneously using hemagglutination-inhibition (HAI) antibody assay for Influenza H1N1, H3N2, and Influenza B strains. Peripheral blood RNA (PaxGene) obtained from blood specimens at each visit were analyzed using Illumina Human HT-12 v4. The study was repeated 2010-2011.

**GSE68004:** Jaggi *et al.* (Jaggi et al., 2018) prospectively enrolled and obtained blood samples in a total of 162 children < 18 years of age at three hospitals (Nationwide Children's Hospital in Columbus, OH, Children's Medical Center in Dallas, TX and Rady Children's Hospital in San Diego, CA) hospitalized with Kawasaki Disease or other febrile illnesses and 37 healthy, asymptomatic children as controls who were age-, gender- and race-matched. Of the 125 hospitalized children, 19 patients had confirmed adenovirus infection and only these samples and the controls were used for our analysis. Healthy controls were enrolled when undergoing elective surgery for non-infectious related causes, or at routine outpatient visits.

**GSE67059:** Heinonen *et al.* (Heinonen et al., 2016) prospectively enrolled previously healthy children <2 years old recruited over six respiratory seasons (2007–2011 and 2013–2015) from four study sites (Columbus, OH; Dallas, TX; Turku, Finland; and Malaga, Spain). Children with respiratory symptoms were enrolled at the emergency department (outpatients) or within 48 hours of hospitalization (inpatients) and were classified into:

HRV- healthy controls (HC; n=37); HRV+ asymptomatic (n=14); HRV+ outpatients (n=30); and HRV+ inpatients (n=70). Exclusion criteria: documented bacterial or viral coinfections, systemic corticosteroid treatment in the preceding 2 weeks, prematurity (<36 wk of gestation), immunodeficiency, or chronic medical conditions. Whole blood RNA samples were hybridized into Illumina arrays. Healthy control subjects were enrolled during well-child visits or minor elective surgical procedures not involving the respiratory tract who did not have a history of a respiratory tract infection within 2 weeks of enrollment. Blood and nasopharyngeal (NP) samples were collected at enrollment from all subjects and analyzed for transcriptional profiles and respiratory viruses. Disease severity was assessed according to the need for hospitalization (inpatients [moderate or severe disease] vs. outpatients [mild disease]) and using a standardized clinical disease severity score (CDSS).

**GSE66099:** Sweeney *et al.* (Sweeney et al., 2015) created this dataset which is composed of unique patients present in the six other GEO datasets published by the Genomics of Pediatric SIRS and Septic Shock Investigators, which includes all unique patients from GSE4607 (described below), GSE8121 (Shanley et al., 2007), GSE9692 (Cvijanovich et al., 2008), GSE13904 (Wong et al., 2009a), GSE26378 (Wynn et al., 2011), and GSE26440 (Wong et al., 2009b). Only unique patients from the Day 1 timepoint are included. The original studies examined pediatric patients admitted to the ICU, who were later classified as either SIRS (non-infectious) or Sepsis or Septic Shock (infectious). There is also a group of healthy controls. Although the original studies examine patients at both ICU day 1 and ICU day 3, patients here are only aggregated ICU day 1 patients. All samples were downloaded as CEL files and re-normalized with gcRMA using R package 'affy'.

**GSE4607:** Wong *et al.* (Wong et al., 2012; 2008; 2007) enrolled children < 10 years of age admitted to the pediatric intensive care unit (PICU) who met criteria for pediatric-specific septic shock as defined by the 2005 International pediatric sepsis consensus conference. Control patients were recruited from the participating institutions using the following exclusion criteria: any acute illness, a recent febrile illness (within 2 weeks), recent use of anti-inflammatory medications (within 2 weeks), or any history of chronic or acute disease associated with inflammation. Whole blood samples were obtained within 24 hours of admission to the PICU, referred to as "Day 1" of septic shock, for microarray analysis with Affymetrix Human Genome U133 Plus 2.0 GeneChip. All study patients were followed for 28 days to determine survival. A subset of patients with influenza, rotavirus, and varicella, and controls were used in our analysis, and we excluded samples that overlapped with those in GSE66099.

**GSE6269:** Ramilo *et al.* (Ramilo et al., 2007) profiled PBMCs from patients (age ≤ 18 years) with acute infections caused by: (1) an RNA virus (influenza A); (2) two gram-positive bacteria (*Staphylococcus aureus* and *Streptococcus pneumoniae*); and (4) a Gram- negative bacterium (*Escherichia coli*). Patients were treated according to standard hospital protocols, and antimicrobial therapy was promptly initiated in the emergency department. The patients were treated with up to three different drugs from an overall set of 13 drugs. The study profiled these samples using three types of microarrays from two manufacturers, Affymetrix (HG U133A and HG U133 plus 2.0) and Illumina, representing technical variability.

**GSE61821:** Hoang *et al.* (Hoang et al., 2014) recruited patients from two cohorts 1) Mild influenza and other febrile illness (OFI) samples, and 2) moderate and severe samples as described below.

**Mild and OFI cohort:** Patients were recruited between January 2008 and January 2010 from an undifferentiated fever inclusion study (EDEN) in Singapore, who were ≥18 years of age and presented ≤72 h from onset of fever ≥38°C. Patients were tested by PCR for influenza A and B, RSV, parainfluenza 1–3, coronavirus, metapneumovirus, enterovirus and adenovirus in nasal swabs and for dengue virus 1–4, human parvovirus B19, Cytomegalovirus, and Epstein Barr virus in EDTA blood. They performed whole-blood transcriptional profiling on all samples from both groups at 72 hours after fever onset, at 3–8 days and 3–4 weeks after self-reported fever onset.

**Moderate and severe cohort:** Samples of moderate and severe influenza patients were collected from a multi-center, double-blinded, randomized control trial of different doses of oseltamivir for the treatment of hospitalized patients with influenza (Registered ClinicalTrials.gov: NCT00298233). The study took place across 13 hospitals in the Southeast Asia Infectious Disease Clinical Research Network (SEAICRN). Whole-blood samples collected before oseltamivir treatment (study day 0) and at follow up (day 28) were used. The inclusion criteria were: (i) age ≥1 year, (ii) duration of illness ≤10 (non-H5N1) or ≤14 (H5N1) days, (iii) positive result for influenza virus A or B using a rapid antigen test or qualitative RT-PCR in a respiratory specimen, (iv) presence of at least one respiratory symptom (cough, dyspnea or sore throat), (v) disease requiring hospital admission, and (vi) one of the following signs of severe influenza: (a) new infiltrate on a chest X-ray, (b) tachypnea (respiratory rate ≥30 for ages ≥12 years, rate ≥40 for ages 6 to 12 years, rate ≥45 for ages 3 to 6 years, rate ≥50 for ages 1– to 3 years), (c) dyspnea (unable to speak full sentences, or use of accessory respiratory muscles), or (d) hypoxia (arterial oxygen saturation ≤92% on room air by a transcutaneous method). Subjects infected with avian influenza virus A/H5N1 were enrolled with any degree of severity. The exclusion criteria were: (i) pregnancy or urine β-hCG positivity, (ii) breast feeding, (iii) prior oseltamivir therapy for >72 hours duration or double dose (any duration) within the past 14 days, (iv) allergy or severe intolerance of oseltamivir, (v) creatinine clearance (CrCl) <10 mL/min. Severe influenza was defined as: i) requiring mechanical ventilation or ii) presenting with severe tachypnea (respiratory rate ≥30 for ages ≥12 years, rate ≥40 for ages 6 to 12 years, rate ≥45 for ages 3 to 6 years, rate ≥50 for ages 1–to 3 years) and hypoxia (arterial oxygen saturation ≤92% on room air by a transcutaneous method. Gene expression microarray was performed using one-color array technology on the Illumina platform (Illumina Inc, San Diego, CA, USA).

**GSE61754:** Davenport *et al.* (Davenport et al., 2014) recruited 22 healthy volunteers aged 18–45 years, and 11 were vaccinated with $1.5\times10^{8}$ plaque forming units (pfu) of MVA-NP+M1. Thirty days after vaccination, all volunteers underwent intranasal challenge with H3N2 influenza (A/Wisconsin/67/2005) at a dose of 1 ml of $10^{5.25}$ TCID50/ml in a quarantine facility. Volunteers were all challenged within a 2-h period. All subjects had hemagglutination inhibition (HI) titers of <1:10 to the challenge virus on admission to the quarantine unit. After viral challenge, volunteers had a physical examination by a trial physician daily and self-reported their symptoms using a modified Jackson scoring system (Zaas et al., 2009) twice daily, from 12-h post-challenge, for 6 days. This system lists upper respiratory and systemic symptoms on a scale of 0–3: 'no symptoms', 'just noticeable', 'bothersome but can still do activities' and 'bothersome and cannot do daily activities.' Scores were summed over the duration of the challenge period to give a total symptom score. Those volunteers with scores of ≥4 and positive viral culture from nasal wash samples were categorized as having laboratory-confirmed influenza (LCI). Severity of infection was graded as mild if the summed symptom score was 4–28, with scores of ≥29 classified as moderate/severe. Volunteers were kept in quarantine until day 6 post-challenge, and all had a negative rapid antigen test on nasal wash sample prior to discharge. Whole blood was collected in PAXgene tubes prior to influenza challenge (day 30) and then at three further time points (12, 24 and 48 h post-challenge) for genome-wide gene expression analysis using the Illumina HumanHT-12 v4 Expression BeadChip. All nasal washing and blood collections for viral shedding and gene expression analysis were taken in the same order that the volunteers were challenged in.

**GSE40012:** Parnell *et al.* (Parnell et al., 2012) recruited patients with severe community-acquired pneumonia requiring ICU admission, patients with noninfective systemic inflammatory response syndrome (SIRS), and healthy controls. They defined SIRS as the presence of at least two of the following four clinical criteria: (a) fever or hypothermia (temperature > 100.4°F (38°C) or < 96.8°F (36°C)); (b) tachycardia (> 90 beats/min), (c) tachypnea (> 20 breaths/min or PaCO2 < 4.3 kPa (32 mm Hg)), or the need for mechanical ventilation; (d) an altered white blood cell count of > 12,000 cells/µl, < 4,000 cells/µl, or the presence of > 10% band forms. Pneumonia was defined as a microbiologically confirmed infection of the lungs in a patient fulfilling the SIRS criteria. Patients were between ages 22 to 75 years. Influenza A H1N1 2009 was confirmed by PCR, and bacterial pneumonia by microbiological cultures. The first blood sample from each patient was collected within 24 hours of ICU admission. Patients were monitored for up to 5 days for microarray analysis of longitudinal gene-expression profiles.

**GSE38900:** Mejias *et al* (Mejias et al., 2013) prospectively enrolled children over six respiratory seasons including a cohort of hospitalized infants (age <2 years) with RSV, HRV, and influenza infections from three centers: (1) Nationwide Children's hospital, Columbus, Ohio, (2) Turku University (Turku, Finland), and (3) Children's Medical Center, Dallas, Texas and blood samples were collected for microarray analysis. Children with documented viral or bacterial co-infections (bacteremia, urinary tract infection, meningitis, acute gastroenteritis, or any bacterial pathogen isolated from a sterile site) were excluded. Children with congenital heart disease, chronic lung disease, immunodeficiency, prematurity (<36 weeks), and systemic steroid treatment within 2 weeks prior to presentation were also excluded. Control samples were obtained from healthy children without comorbidities, use of systemic steroids, or presence of any illness within 2 weeks prior to enrollment who were either undergoing elective surgery not involving the respiratory tract, or at outpatient visits for routine care. To exclude viral co-infections, respiratory samples were tested by viral culture or PCR in 94% of patients and controls.

**GSE2729:** Wang *et al.* (Wang et al., 2007) collected blood and fecal specimens from children < 3 years of age who were hospitalized for treatment of acute rotavirus gastroenteritis, who were otherwise generally in good health with no prior history of rotavirus diarrhea and had not received rotavirus vaccines. All patients had symptoms typical of rotavirus infection (fever, vomiting, diarrhea, or dehydration), and all were confirmed positive by the testing of fecal specimens by using enzyme immunoassay and PCR. Blood samples were drawn at the time of hospital admission, within 72 hours of enrollment, and 3 weeks later from some children. PBMCs were isolated for microarray analysis using Affymetrix Human U95Av2 high density oligonucleotide arrays. Controls were age-matched healthy children admitted for elective surgery with no symptoms of gastroenteritis and fecal specimens that tested negative for rotavirus by enzyme immunoassay.

**GSE27131:** Berdal *et al.* (Berdal et al., 2011) prospectively recruited ICU patients over a four week period who were > 18 years of age diagnosed with pandemic H1N1 influenza by upper airway PCR with bilateral chest X-rays infiltrates requiring mechanical ventilator support. Patients with significant comorbidity (malignancy and severe systemic inflammatory diseases) were excluded. Age and gender-matched healthy individuals served as controls. Peripheral venous blood was collected for microarray analysis at inclusion and after 3 and 6 days. Blood was centrifuged at 2500× *g* for 20 min to obtain platelet-poor plasma (EDTA) or allowed to clot before centrifugation, after which RNA from blood leukocytes was obtained by PAXgene Blood RNA System for microarray analysis.

**GSE25504:** Smith *et al.* (Smith et al., 2014) and Dickinson *et al.* (Dickinson et al., 2015) enrolled infants from the Neonatal Unit, Royal Infirmary of Edinburgh, and the Division of Pathway Medicine, University of Edinburgh having blood cultures taken for suspected infection. They obtained whole blood for expression profiling at the time of first clinical signs of suspected and microbiological blood culture. The infected group was defined as patients with suspected clinical infection that proved to have microbiological evidence of infection from a usually sterile body site, and samples were only included as positive if both clinicians agreed that infection was present. Exclusion criteria were: infants of mothers known to be positive for hepatitis B, HIV or hepatitis C viruses, mothers with known history of drug misuse, and mothers that had no antenatal screening for blood-borne viruses. Other exclusion criteria were infants who did not require clinical blood samples and infants for whom extra blood sampling might be of particular risk (such as anemia). Controls were defined as 'well' infants having blood taken for other clinical reasons, which included screening tests for maternal thyroid disease, routine neonatal screening, pigmented scrotum, electrolyte check for previously deranged sodium, bilirubin check due to jaundice, and neonatal encephalopathy, all of which were normal. Samples were divided into four cohorts which were each run on different microarray platforms, three of which were excluded from our meta-analysis (Affymetrix Human Genome U133 Plus 2.0, Illumina HumanHT-12 V3.0 expression beadchip, and

Codelink 55K Human Array) because they did not have at least 2 samples present in each category (viral infection vs control). Patient samples with bacterial or fungal infection were also excluded from our analysis. We therefore only analyzed samples from 9 infants with corrected gestational age ranges of $27^5$ to $39^6$ weeks and ran on Affymetrix Human Genome U219 Array from this dataset, which included 6 controls and 3 patients with viral infections (2 Rhinovirus, 1 CMV).

**GSE21802:** Bermejo-Martin *et al.* (Bermejo-Martin et al., 2010) recruited critically ill patients aged 18 to 65 years with influenza pneumonia during the acute phase of illness with acute respiratory distress and unequivocal alveolar opacification involving two or more lobes with negative respiratory and blood bacterial cultures on admission to the ICU. Only patients with confirmed H1N1 infection by RT-PCR were included in the study. Age-matched healthy volunteers were also recruited between workers of the University of Valladolid, Spain. Treatment decisions for all patients, including corticosteroid therapy, were not standardized and were used at the discretion of the attending physician.

**GSE20346:** Parnell *et al.* (Parnell et al., 2011) recruited hospitalized patients aged 21-75 years with severe infection, defined as the presence of at least one major organ failure requiring critical care intervention. These patients had either viral (seasonal H3N2 or pandemic H1N1/09 influenza virus) or bacterial infections and were followed for four days. Healthy volunteers were enrolled from a local influenza vaccination program.

**GSE17156:** Zaas *et al.* (Zaas et al., 2009) inoculated healthy volunteers (HV) with one of the following viruses (HRV, RSV, influenza) as described below.

**HRV cohort:** The HRV challenge subjects were recruited through an active screening protocol at the University of Virginia (Charlottesville, VA). On the day of inoculation, $10^6$TCID50 GMP HRV serotype 39 was administered intranasally. Subjects were admitted to a quarantine facility for 48 hours post-inoculation. Nasopharyngeal (NP) lavage samples were obtained from each subject daily for titers to accurately gauge the success and timing of inoculation. After 48 hours post-inoculation, subjects were released from quarantine and returned for 3 consecutive days for further sample acquisition.

**RSV cohort:** The RSV challenge was performed by Retroscreen Virology, Ltd. (London). On the day of inoculation, a dose of $10^4$ TCID50 RSV (serotype A) was administered intranasally. Blood and NP lavage samples were collected as in the HRV cohort, but continued throughout quarantine. Due to the incubation period of RSV A, subjects were not released from quarantine until they were 288 hours post-inoculation and negative for rapid RSV antigen detection.

**Influenza cohort:** The influenza challenge A/Wisconsin/67/2005 (H3N2) was performed at Retroscreen Virology, Ltd. (Brentwood, UK). On the day of inoculation, a $10^6$ dose of TCID50 influenza A was diluted and administered intranasally per standard methods at different doses: (1:10, 1:100, 1:1000, 1:10000) with 4-5 subjects receiving each dose. Due to the incubation period, subjects were not released from quarantine until 168th hours post-inoculation. Blood and NP lavage samples were collected throughout the duration of quarantine. All subjects received oral oseltamivir (Roche Pharmaceuticals) (75 mg) by mouth twice daily at day 6 post-inoculation and were negative by rapid antigen detection at the time of discharge.

**GSE117827:** Yu *et al.* (Yu et al., 2019) profiled children age 3 months to 18 years hospitalized for acute respiratory illness with positive results only for a single virus (Rhinovirus, RSV, Enterovirus, Coxsackievirus) on a multiplex nasopharyngeal swab obtained as part of the child's routine care (BioFire FilmArray Respiratory Panel, Salt Lake City, UT). Exclusion criteria included any underlying medical condition that required regular medical care, receipt of immunosuppressive medications, corticosteroids within the preceding 30 days, and receipt of antibiotics within 7 days. Controls were age-matched children having ambulatory surgery for non-acute conditions. Exclusion criteria were the same as for symptomatic subjects plus fever within the preceding 48 hours. Blood samples were profiled using microarray with Affymetrix.

**GSE111368:** Dunning *et al*. (Dunning et al., 2018) and the Mechanisms of Severe Acute Influenza Consortium (MOSAIC) profiled whole blood for microarray analysis from patients aged ≥ 16 years during two successive winters (2009–2010 and 2010–2011) with pandemic H1N1 and seasonal H3N2 (determined by RT-PCR) admitted to general wards or ICU. They included patients with prior or concurrent comorbidities: (most commonly asthma, pregnancy, immunocompromising conditions or co-infection with other respiratory pathogens), to reflect the populations known to be at greatest risk of severe influenza. Adult healthy age, sex and ethnicity-matched control subjects screened to exclude known illnesses or current use of medications were also included (Registered Clinical Trial NCT00965354). Samples were obtained at three time points: T1 (recruitment), T2 (approximately 48 h after T1) and T3 (at least 4 weeks after T1). Severity of illness was graded as: 1, no substantial respiratory compromise, with blood oxygen saturation of > 93% while the patient was breathing room air; 2, oxygen saturation of ≤ 93% while the patient was breathing room air, justifying or requiring supplemental oxygen by face mask or nasal cannulae (with or without continuous positive airway pressure support or non-invasive mechanical ventilation); 3, respiratory compromise requiring invasive mechanical ventilation with or without ECMO (extracorporeal membrane oxygenation). Nasopharyngeal aspirates and swabs collected at T1 underwent microscopy and culture for bacteria, and multiplex PCR was performed to detect S. aureus, C. pneumoniae, H. influenzae, S.pneumoniae, Pneumocystis pneumoniae, Legionella species, Klebsiella pneumoniae, Salmonella species, Moraxella catarrhalis, Mycoplasma pneumoniae and Bordetella pertussis. Throat swab samples obtained at T1 also underwent culture and microscopy. Where available, urine samples collected between T1 and T2 underwent pneumococcal antigen testing (BinaxNow, Alere). An independent microbiologist assessed the significance and validity of positive blood-culture results, in an attempt to exclude cases of pseudobacteremia.

**GSE103842:** Rodriguez-Fernandez *et al.* (Rodriguez-Fernandez et al., 2017) prospectively recruited previously healthy infants (age < 2 years) hospitalized to general wards or Pediatric ICU over 5 nonconsecutive respiratory seasons with RSV bronchiolitis. Patients were excluded if they were premature (gestational age ≤36 weeks), had a previous episode of documented RSV infection, chronic medical conditions, immunodeficiency, or had received systemic steroids or immunomodulatory drugs within 2 weeks of hospitalization. Healthy asymptomatic controls were enrolled during well-child visits or minor elective surgical procedures not involving the respiratory tract. Bronchiolitis was defined as the presence of rhinitis, tachypnea, wheezing, cough, crackles, use of accessory muscles, and/or nasal flaring with or without fever. Children who met the inclusion criteria and had a positive RSV test per standard of care, were enrolled within 24 hours of hospitalization, and a nasal wash sample was obtained for RSV typing and viral load quantitation. Nasal wash samples were also tested for other respiratory viruses in 75% of patients; 97% using a multiplex assay targeting 17 pathogens and 3% by viral culture. In a subset of infants, blood samples (1–3 mL) were collected and blood RNA was processed and hybridized into Illumina Human HT12 V4 beadchips and scanned on the Illumina Beadstation 500.

**GSE101702:** Tang *et al.* (Tang et al., 2019) profiled blood samples from hospitalized patients with varying severity of influenza infection. The eligibility criteria included (1) age > 18 years and (2) World Health Organization definition of influenza-like illness (fever of 38 C° or higher, cough and illness onset within the last ten days), and patients with a high likelihood of infection, based on history and clinical features. Airway samples (nasopharyngeal swab, throat sample or sputum) and a 2.5ml peripheral blood sample (into PAXgene tubes) were obtained in each participant within 24 hours of their presentation to the hospital. Airway samples were tested for bacterial pathogens and common respiratory viruses. Blood samples in PAXgene tubes were later processed for microarray analysis (Agilent 8× 60k Human V3) of participants and healthy control subjects without any medical illnesses. In patients admitted to the intensive care unit (ICU), additional respiratory samples were obtained from bronchoalveolar lavage or tracheal aspirates. Standard microbiological testing was performed in these samples, including sputum Gram stain and culture. Testing for atypical respiratory pathogens (Chlamydophila pneumoniae, Mycoplasma pneumoniae, and Legionella pneumophila) was performed in selected patients at the discretion of treating physicians. All patients were tested for respiratory viruses using nucleic acid PCR. The PCR panel included primers for influenza A, influenza B, respiratory syncytial virus, rhinovirus, parainfluenza virus, and human metapneumovirus. Patients infected with other viruses than influenza were excluded.

**E-MTAB-5195:** Jong *et al.* (Jong et al., 2016) studied respiratory syncytial virus (RSV) infection in 39 hospitalized young children (age < 2 years) using early blood microarray transcriptome profiles followed until recovery and of which the level of disease severity was determined retrospectively. Patients were recruited from three hospitals, and nasopharyngeal wash and blood samples were prospectively obtained from patients with a viral bronchiolitis. Samples were taken within 24 hours after first contact with the hospital, and only patients with an RSV infection, as retrospectively determined by PCR were included in the study. Exclusion criteria were: immunodeficiency, systemic steroid treatment in the previous 2 weeks, blood transfusion, congenital heart and chronic lung disease. Patients were followed until recovery and were retrospectively classified as: mild for children without hypoxia, moderate for patients requiring supplemental oxygen (oxygen saturations < 90%, ≥ 10 minutes) and severe for children requiring mechanical ventilation due to apnea, exhaustion and/ or respiratory failure. Recovery samples were obtained after 4–6 weeks, during home visits. Blood samples were obtained from healthy controls without underlying diseases or medication subjected to elective surgery.

Zhai, Y., Franco, L.M., Atmar, R.L., Quarles, J.M., Arden, N., Bucasas, K.L., Wells, J.M., Nino, D., Wang, X., Zapata, G.E., et al. Host transcriptional response to influenza and other acute respiratory viral infections - a prospective cohort study. PLoS Pathog *11*, e1004869.
